# The effects of communicating uncertainty around statistics on public trust: an international study

**DOI:** 10.1101/2021.09.27.21264202

**Authors:** John R. Kerr, Anne Marthe van der Bles, Claudia Schneider, Sarah Dryhurst, Vivien Chopurian, Alexandra L.J. Freeman, Sander van der Linden

## Abstract

A growing body of research indicates that transparent communication of statistical uncertainty around facts and figures does not undermine credibility. However, the extent to which these findings apply in the context of the COVID-19 pandemic—rife with uncertainties—is unclear. In a large international survey experiment, (Study 1; *N* = 10,519) we report that communicating uncertainty around COVID-19 statistics in the form of a numeric range (vs. no uncertainty) may lead to slightly lower trust in the number presented but has no impact on trust in the source of the information. We also report the minimal impact of numeric uncertainty on trust is consistent across estimates of current or future COVID-19 statistics (Study 2) and figures relating to environmental or economic research, rather than the pandemic (Study 3). Conversely, we find imprecise statements about the mere existence of uncertainty without quantification can undermine both trust in the numbers and their source – though effects vary across countries and contexts. Communicators can be transparent about statistical uncertainty without concerns about undermining perceptions of their trustworthiness, but ideally should aim to use numerical ranges rather than verbal statements.

---

How much can a specific number, projection or claim be trusted or weighted in a decision? In some cases uncertainty can be quantified and communicated in numeric terms, for example a 95% confidence interval (e.g. ‘UK unemployment fell by 116,000 (range between 17,000 and 215,000)’), or as a visual representation of the quantified uncertainty such as an error bar around a plotted point estimate. At the other end of a spectrum of precision, uncertainty can simply be acknowledged as merely existing with vague statements such as ‘there is uncertainty around the exact figure—it could be higher or lower’ (for a more comprehensive taxonomy of how uncertainty can be represented, see (van der Bles et al., 2019)).

In many situations, however, such uncertainties are omitted by individuals or organization when communicating information to the public (e.g., researchers, government organisations). This may be due to a sense that uncertainty information might undermine audiences’ trust in the information or its source more than is warranted, make content less easy to understand, bias perceptions, evoke negative emotions, or reduce decision making quality (Fischhoff, 2012; Hullman, 2020; Manski, 2020; Post & Maier, 2016; van der Bles et al., 2020).

Previous research has investigated how different ways of communicating uncertainty affect audiences’ perceptions of credibility—both in terms of how accurate or reliable the information is perceived to be and in terms of more general trust in, or perceived expertise of, those doing the communicating. For example, van der Bles et al. (2020) reports that including uncertainty information in the form of a numeric range (compared to a specific point estimate) has little impact on readers’ trust in the information or its source. These findings are reflected in the broader literature: numerical quantification of uncertainty as a range or confidence interval has little to no negative effect on credibility and indeed in some cases may increase credibility (Gustafson & Rice, 2019, 2020; Han et al., 2011; Joslyn & Demnitz, 2019; Joslyn & LeClerc, 2016; Lipkus et al., 2001; McDowell & Kause, 2021).

Verbal communication of imprecise, unquantified uncertainty however, can undermine the credibility of a message and its source. van der Bles et al (2020) also find that the inclusion of sentences stating that there is some uncertainty around a reported figure and the actual value “could be somewhat higher or lower” consistently resulted in lower trust in the information and in some cases lower trust in the source of the information. Research investigating other forms of verbal, unquantified uncertainty provides mixed results. For example Thiebach et al. (2015) report that the presence or absence of hedge words such as ‘may’ and ‘possibly’ has no bearing on the perceived credibility of statements. Han et al., (2018) reports that presence of vague statements describing uncertainty around influenza severity and vaccine efficacy (‘…we are not sure exactly how effective it will be…’) in a news release undermines trust in the primary source. Conversely, Nakayachi et al. (2018) find that coupling earthquake risk estimates with a statement explaining assessments come with ‘high uncertainty’ increased readers’ trust in seismologists, though this effect was only detected when low (20% vs 70% or 100%) quake probabilities were communicated.

This research, however, has been conducted only within a limited range of samples from WEIRD countries (Henrich et al., 2010), and on topics that may not have had immediate emotional salience to the audience (despite efforts to explore some topical and divisive issues such as climate change and immigration). In the current research we build and expand on the findings of van der Bles et al. (2020), examining how inclusion of uncertainty information, expressed as a numeric range or verbal statement, impacts readers’ trust in a COVID-19 related statistics and their source. We also investigate how these effects vary across different national contexts.

At the time of data collection for the current research (Studies 1 and 2) it is not an exaggeration to say that the world was preoccupied with COVID-19; citizens of many countries were under wide-ranging restrictions (Hale et al., 2021), news of the virus and its impact dominated mainstream and social media (Pearman et al., 2021; Tsao et al., 2021), and the perceived risk from the virus was high across many countries (Dryhurst, Schneider, et al., 2020; Schneider et al., 2021).

At the same time, the SARS-CoV2 pandemic brought uncertainties of all kinds to a broad range of people: uncertainty about the potential future course of the pandemic and rate of transmission; uncertainty about the current number of cases, or deaths, or mortality rate; uncertainty about the underlying disease course and risk factors associated with a poor outcome (Koffman et al., 2020). Inherent in communication of facts about the disease and the pandemic, then, is the need to communicate this uncertainty in a way that allows people to understand it and make appropriate judgements about how much to trust it and weight the information in their decision-making given its level of certainty.

Calls have been made for transparent, trustworthy communication of uncertainties around COVID-19 (Rutter et al., 2020; Veit et al., 2020). Yet many official communications on the topic do not explicitly acknowledge such uncertainties, possibly due to concerns over trust and credibility. Notably, one experiment examining the impact of uncertainty on public trust in science suggests these concerns are not unfounded. Kreps and Kriner (2020) report that participants presented with an estimate of future COVID-19 deaths expressed less general trust in scientists when that estimate was provided as a (relatively large) range rather than a single number. The authors conclude that expressing uncertainty as a range: “may be more intellectually honest, but it nonetheless comes at a cost of eroding public confidence.” (p. 5) This worrying conclusion presents a challenge to calls for transparent communication as well as conflicting with much of the prior research on this matter. Indeed, a recent systematic review of uncertainty communication experiments found no studies in which communication of technical (i.e. statistical) uncertainty had a negative effect on credibility of either the claim or its source (Gustafson & Rice, 2020).

In the current studies, we seek to add to the limited empirical literature on how uncertainty around COVID-19 information is received. We examine how the presence and format of uncertainty in communications impacts perceptions of uncertainty, and message and source credibility. We first carried out an experimental study in 12 countries across Europe, North and Central America, and Asia (Study 1), conducted between mid-March and mid-April 2020, in which participants were randomised to one of three conditions: a control condition of receiving COVID-19 statistics with no uncertainty (a point estimate), a condition where the information included a numerical range around the estimate, and a condition in which the uncertainty was not quantified but communicated as an imprecise verbal statement.

The uncertainty in this study related to the COVID-19 infection hospitalisation rate – that is, the proportion of people infected with COVID-19 who require hospitalisation for treatment of symptoms. As such, the uncertainty can be considered epistemic, arising from a lack of data or knowledge. In this sense we use epistemic uncertainty to refer to uncertainty regarding past or current phenomena; unknowns which in theory could be known (van der Bles et al., 2019). Some often-reported COVID-19 statistics are imbued with another type of uncertainty - aleatory uncertainty—meaning that they simply cannot be known for certain due to the inherent indeterminacy or randomness of the world - such as predicted future numbers of cases or deaths (van der Bles et al., 2019). A number of uncertainty communication studies have examined how people perceive and interpret verbal and numeric expressions of uncertainty relating to future risks and events (e.g., Joslyn & LeClerc, 2016; Nakayachi et al., 2018). However we are not aware of any research which has systematically examined how the impact of uncertainty format (e.g. numeric or verbal) on trust varies depending on whether the information provided relates to the present (epistemic) or future (aleatory). In Study 2, we therefore explore potential differences in the impact of these two types of uncertainty by including a further experimental factor to the Study 1 design: varying whether COVID-19 death statistics presented refer to current estimates or future forecasts.

Finally, to further examine the generalisability of our findings to other domains, which participants may perceive as more psychologically distant, we also present results from a third experiment (Study 3). This study investigates how numeric or verbal uncertainty, presented as either epistemic or aleatory, is perceived when it relates to environmental or economic issues, rather than COVID-19.

## Study 1

### Methods

Participants were recruited across 12 countries. Participants in Australia were recruited through Dynata (dynata.com), participants in France were recruited by BVA (bva-group.com) and US and UK samples were recruited via Prolific (prolific.ac). Participants in all other countries were recruited through Respondi (respondi.com). Data collection was carried out between March and May, 2020, with each survey fielded for approximately five days. Interlocking quotas were used to match samples to the national profile on age and sex (and ethnicity for Prolific samples). Sample sizes and characteristics are reported in table 1. Surveys were translated from English to other languages by native speakers fluent in English.

**Table 1:**
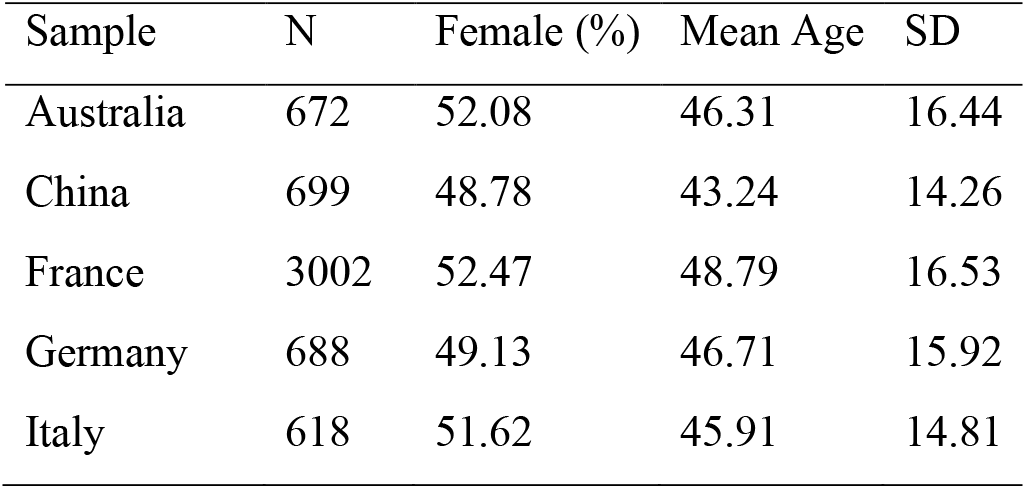
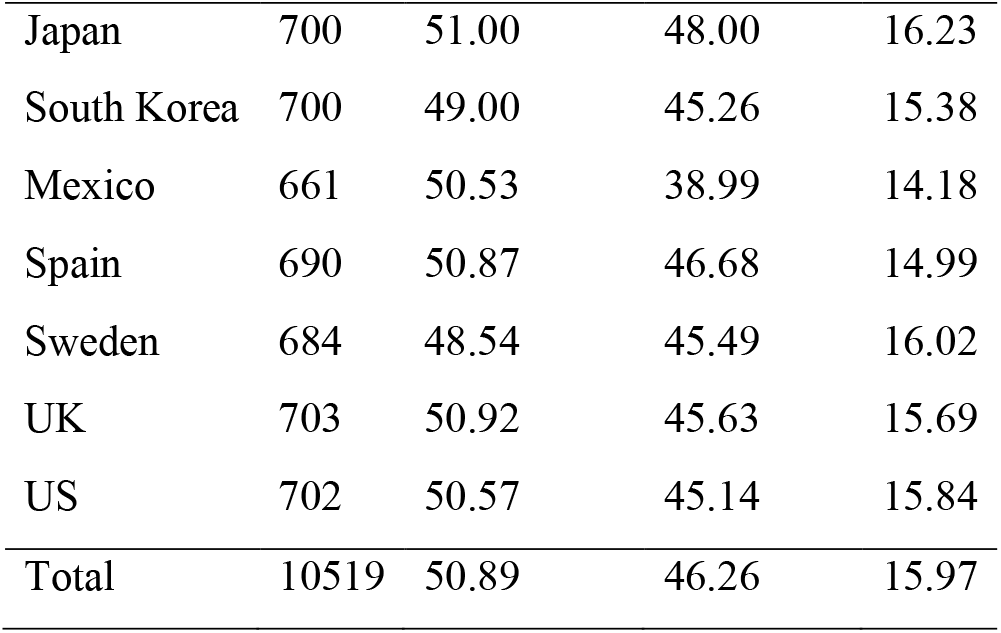
Samples included in study.

The study was approved by the University of Cambridge’s Psychology Research Ethics Committee (PRE.2020.034).

A GPower (Erdfelder et al., 1996) power calculation indicated that sample sizes were sufficient to detect effect sizes reported in previous research, e.g., a *d* = .55 effect of verbal uncertainty (vs control) on trust in numbers, (see internal meta-analysis, van der Bles et al., 2020), at 95% power and an alpha level of 0.05.

Participants completed an online survey experiment embedded in a larger survey including questions on COVID-19 risk perceptions and attitudes, hosted on the Qualtrics survey platform. In the survey, participants were randomly assigned to one of three ‘format’ conditions. All participants were shown a piece of text about COVID-19: “*Illness due to COVID-19 infection is generally mild, especially for children and young adults. However, it can cause serious illness: for people aged 70-80, about 17% of those who catch it need hospital care*.*”*^1^ In the control format condition this was the exact text shown. In the ‘numerical uncertainty’ format condition, the text shown to participants additionally had the phrase “*(range between 10% and 34%)*” inserted after the percentage figure. In the ‘verbal uncertainty’ format condition, the text shown to participants included the additional sentence “*There is some uncertainty about that percentage, it could be somewhat higher or lower*.” at the end (but no numerical range was shown).

Participants were then asked a series of questions forming our main dependent variables. Perceived uncertainty was measured with the item “*To what extent do you think this number is certain or uncertain?*” (responses: 1 = ‘*Very certain*’ to 7 = ‘*Very uncertain*’).

Trust in the number was measured as the average of three items: “*To what extent do you think this number is reliable?”, “To what extent do you think this number is accurate?”*, and “*To what extent do you think this number is trustworthy?”* (1 = ‘*Not at all*’ to 7 = ‘*Very*’, Cronbach’s αs .90-.96). Trust in the source of the information was measured with the item “*To what extent do you think that the people responsible for producing this number are trustworthy?” (1 = ‘Not at all’* to 7 = ‘*Very’*). Participants also answered questions about how positive or negative they felt about the information and how easy they found it to read. Details and results for these secondary outcomes are reported in Appendix 1.

### Results

Figure 1 shows the pooled means from all participants on perceived uncertainty, trust in numbers and trust in the source of the numbers across the three experimental format conditions.

**Figure 1:**
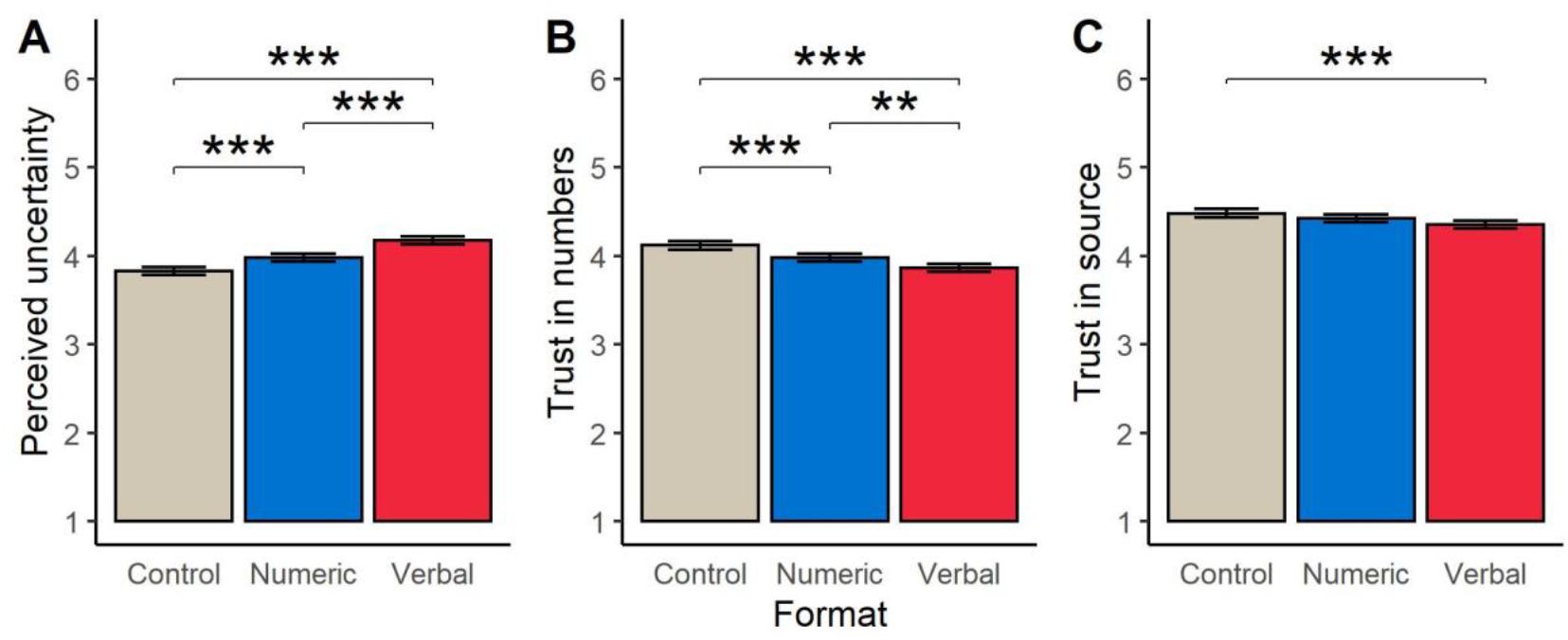
Mean levels of perceived uncertainty (A), trust in number (B) and trust in source (C) across experimental conditions. Whiskers indicate 95% confidence interval; horizontal brackets indicate significant difference between conditions: \*\**p* < .01, \*\*\**p* < .001.

A one-way ANOVA analysing the pooled data from all country samples^2^ revealed a significant effect of experimental condition on *perceived uncertainty, F*(2, 10472) = 56.54, *p* < .001, η^2^ = 0.011. Post hoc analyses (Tukey’s HSD), revealed that participants who read a message with a numeric expression of uncertainty, perceived, on average, more uncertainty than those who read the control message (*M*_control_ = 3.82, *SD* = 1.38; *M*_numeric_ = 3.98, *SD* = 1.35; *p* < .001, Cohen’s *d* = .11). Participants who read the message containing a verbal expression of uncertainty perceived more uncertainty than those who read the numeric uncertainty message (*M*_verbal_ = 4.17, SD = 1.38; *p* < .001, *d* = .14), or the control message (*p* < .001, *d* = .25).

There was also a significant effect of message condition on reported *trust in the numbers* presented, *F*(2, 10454) = 30.50, *p* < .001, η^2^ = 0.006. As detailed in Figure 1B, participants in the numeric uncertainty condition reported a lower level of trust in the numbers compared to those in the control condition (*M*_control_ = 4.12, *SD* = 1.41; *M*_numeric_ = 3.98, *SD* = 1.35; *p* < .001, *d* = .10), and participants in the verbal uncertainty condition reported a lower level of trust in the numbers than those in the numeric uncertainty (*M*_verbal_ = 3.86, *SD* = 1.36; *p* < .01, *d* = .09) and control conditions (*p* < .001, *d* = .18).

Finally, there was a small effect of experimental condition on *trust in the source* of the information, *F*(2, 10467) = 6.97, *p* < .001, η^2^= 0.001 (see Figure 1C). Participants in the verbal uncertainty condition reported a lower level of trust in the source of the information compared to those in the control condition (*M*_control_ = 4.48, *SD* = 1.44; *M*_verbal_ = 4.35, *SD* = 1.42; *p* < .001, *d* = .09). No other significant differences were detected between conditions (*M*_numeric_ = 4.42, *SD* = 1.40).

Additional results focusing on secondary outcomes (difficulty of understanding and emotional response) are reported in the supplementary information (Appendix 1). Briefly, we find that participants rated the control format easiest to understand, followed by the verbal then numeric formats. In terms of emotion, participants in the verbal uncertainty condition reported feeling slightly more positive about the information compared to participants in the control condition.

To investigate the results at the country-level, we ran individual one-way ANOVAs examining the effect of experimental condition on each of the three outcome variables, for each country. We summarise the results in Figure 2, with full details presented in Appendix 2.

**Figure 2:**
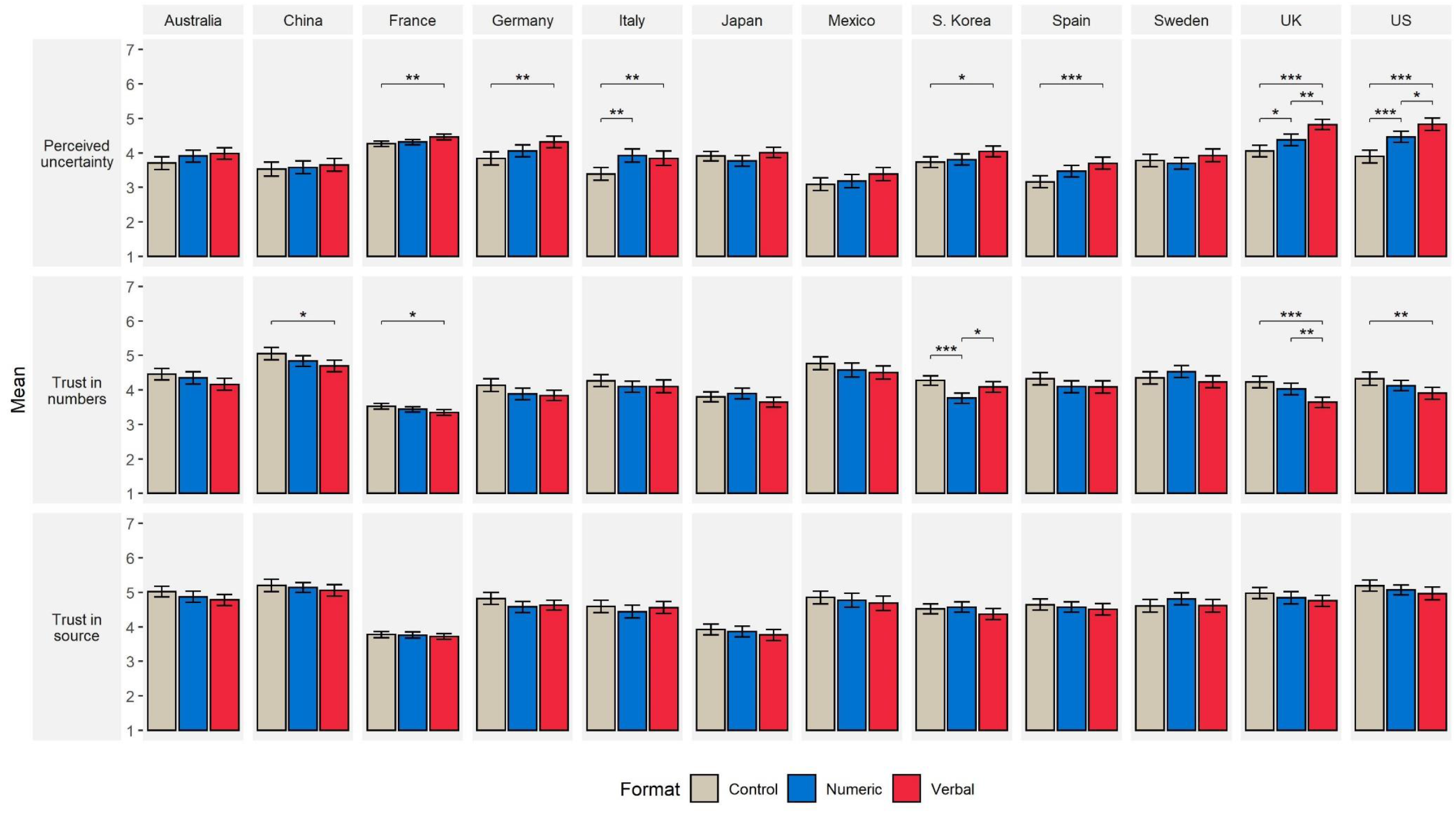
Effect of experimental condition on key outcome variables, across countries. Bars indicate mean and 95% confidence interval. Horizontal brackets indicate significant pairwise difference between conditions (corrected for multiple testing; Benjamini-Hochberg correction applied across all tests). \**p* < .05, \*\**p* < .01, \**p* < .001.

The results of ANOVAs indicated that uncertainty format had a significant effect on perceived uncertainty in samples collected in France, Germany, Italy, South Korea, Spain, the UK and the US. The results of pairwise comparisons between conditions, for each country are presented in Figure 2. Due to the large number of comparisons, we applied a Benjamini-Hochberg correction across all tests to correct for multiple testing. Results revealed a pattern of effects consistent with those reported for the combined sample. Where significant differences in perceived uncertainty were detected between groups, we find that uncertainty is highest in the verbal uncertainty condition, followed by the numeric uncertainty condition and then the control condition. This pattern appears most clearly in our US and UK samples.

Experimental condition also had a significant effect on trust in the numbers in samples collected in China, France, Germany, South Korea, the US, and the UK. Considering pairwise comparisons between conditions, we find the inverse pattern to perceptions of uncertainty where significant differences were detected; reported trust was lowest in the verbal uncertainty condition, followed by the numeric uncertainty condition, then the control condition. There was one exception: in the South Korea sample, participants in the verbal condition reported, on average, a significantly higher level of trust in the numbers compared to those in the numeric condition. However, for most countries examined we report no significant differences between any conditions.

Across all individual country samples, we did not find a significant effect of the uncertainty format condition on trust in the source of the information.

Considering the heterogeneity between countries, we conducted a series of exploratory analyses seeking to explain the variation in effect sizes (reported in Appendix 3). These analyses drew on both external country-level data (risk tolerance data drawn from international studies) and additional items included in the surveys (numeracy and social trust). Given the small number data points (12 countries) no firm conclusions can be drawn. However, we found a significant correlation between the mean level of numeracy (Cokely et al., 2012) in a given sample and the effect size of verbal uncertainty on trust in the numbers presented. Country samples with higher numeracy reported lower trust in numbers presented with verbal uncertainty (relative to control).

### Interim Discussion

This large and international dataset, in an emotionally salient context for participants, broadly confirms the findings of our previous work in other contexts (Dryhurst, van der Bles, et al., 2020; van der Bles et al., 2020): when presented with uncertainty information, people perceived greater uncertainty in the reported hospitalization rate, but there was only a small decrease in trust in the numbers and this effect was larger for the verbal uncertainty condition than the numeric. Considering trust in the source of the numbers, we find that participants were less trusting of the source when verbal uncertainty was communicated. However, this small effect (*d* = .09) was only detectable in the pooled sample combining all countries.

Comparing between countries, we find that the US and UK were most sensitive to uncertainty information in terms of its impact on perceptions of uncertainty. In five of the twelve countries surveyed (Australia, China, Japan, Mexico, and Sweden) we find that neither verbal nor numeric expressions of uncertainty had a significant impact on how uncertain participants rated the information. Similarly, at the individual country level, we find that trust in the numbers presented is not consistently affected by uncertainty information; in four countries (China, France, US, UK) we find that statements with verbal uncertainty are considered significantly less credible than those without, and in South Korea numeric uncertainty is rated as less credible. One consistent finding across countries is that uncertainty information did not significantly impact trust in the source of the information in any one country (though we do report a small significant effect in the pooled sample).

This heterogeneity of effects across countries in terms of trust in numbers is noteworthy. Most previous studies of uncertainty communication have drawn on US and UK samples (e.g., Gustafson & Rice, 2019; Joslyn & LeClerc, 2016; van der Bles et al., 2020). The current results suggest that effects of verbal uncertainty information on perceptions of trustworthiness and credibility may not be universal; cultural context may play an important role in how people respond to such information. Researchers and communicators should be aware that uncertainty communication advice and findings from one context may not translate to another. The reasons for such differences remain unclear. The current study did not aim to provide a cross-cultural examination of how uncertainty information is perceived (though we present an exploratory attempt in Appendix 3); further research is required to fully understand the inter-country variation observed.

We find less inter-country variation when considering numeric uncertainty. Here, the results align with a key finding from van der Bles et al. (2020): communication of numeric uncertainty has little impact on trust in the numbers and no effect on trust in the source (though a small significant effect was detected in the pooled sample). Including statistical uncertainty such as confidence intervals in communications does not undermine credibility.

A final consideration for Study 1 is that the uncertainty presented in this study was purely epistemic: it did not involve the uncertainty of an unknown future event. To compare the public’s reaction to aleatoric uncertainty (uncertainty about the future) to their reaction to this epistemic uncertainty, we carried out a further study.

## Study 2

This study aimed to investigate how the effect of uncertainty information on trust differs as function of uncertainty type (epistemic or aleatory). We conducted a survey experiment similar to Study 1 with the addition of a further experimental factor, varying if the numbers communicated related to the present or future.

### Methods

A total of 2,309 UK adult participants (51.4% female; *M*_age_ = 45.2, *SD* = 15.8) were recruited in May 2020 via online panel providers Respondi (*n* = 1150) and Prolific (*n* = 1159). Quotas were set to match the participant pool to the national profile on age, sex, and, for the Prolific sample, ethnicity. The study was approved by the University of Cambridge’s Psychology Research Ethics Committee (PRE.2020.034).

As in Study 1, participants completed an online survey experiment on the Qualtrics platform. Participants were randomised to one of six conditions, in a 3 (uncertainty format) x 2 (uncertainty type) factorial experiment.

The stimuli in this experiment were drawn from statistics reported by modelling from Imperial College London (n.d.). To ensure that the participants were not misled by using factually inaccurate statistics on such an important topic, real modelling figures had to be used. This does introduce a confound to the experimental design as there was necessarily a difference in point estimates and intervals presented to participants in the different uncertainty format conditions. We return to this point in discussing the results.

Those in the epistemic uncertainty condition were given the core information “Looking at the average number of deaths per day over a period of time helps to understand whether the COVID19 epidemic is stabilising in the United Kingdom. This past week, the average number of deaths in the UK was about 555 per day.” Control participants received only this information. Those in the numeric uncertainty format condition were given the extra information “(range between 439 and 698).” Whilst those in the verbal uncertainty format condition were instead given the extra information “There is some uncertainty about this number, it could be somewhat higher or lower.”

Those in the aleatory uncertainty condition were given the information “Looking at the average number of deaths per day over a period of time helps to understand whether the COVID19 epidemic is stabilising in the United Kingdom. This upcoming week, the average number of deaths in the UK is expected to be about 591 per day.” Control participants received only this information. Those in the numerical uncertainty format condition were given the extra information “(range between 371 and 1081).” Whilst those in the verbal uncertainty format condition were instead given the extra information “There is some uncertainty about this number, it could be somewhat higher or lower.”

Participants were then asked the same questions as in Study 1 to ascertain their perception of the uncertainty in the number, their trust in the number (three items; α = .94) and their trust in the people responsible for producing the number. Participants also answered questions about how positive or negative the information made them feel, how easy it was to understand and how competent they thought the source of the information was. Details and results for these secondary outcomes are reported in the supplementary material (Appendix 4).

### Results

Considering first *perceived uncertainty*, a two-way ANOVA examining the effect of uncertainty format (control, numeric, and verbal) and uncertainty type (epistemic and aleatory), revealed a significant interaction, *F*(2, 2299) = 4.01, *p* = 0.02, η _*p*_^2^= 0.003, indicating that the effect of condition differed across uncertainty types. This was followed up with one-way ANOVAs examining the effect of condition in each uncertainty type group. Format had a significant effect on perceived uncertainty among people who read a message with epistemic uncertainty, *F*(2, 1156) = 42.25, *p* < .001, η_*p*_^2^= 0.068, and aleatory uncertainty, *F*(2, 1143) = 15.24, *p* < .001, η _*p*_^2^ = 0.026.

Post hoc tests revealed that, among participants presented with epistemic uncertainty, verbal uncertainty (*M*_verbal_ = 4.54, *SD* = 1.53) was perceived as more uncertain than numeric uncertainty (*M*_numeric_ = 3.70, *SD* = 1.36; *p* < .001, *d* = .58) or no uncertainty (*M*_control_ = 3.72, *SD* = 1.39; *p* < .001, *d* = .55). There was no significant difference between uncertainty ratings in the control and numeric conditions.

A similar pattern was seen among those presented with aleatory uncertainty, though the effects were smaller: verbal uncertainty (*M*_verbal_ = 4.44, *SD* = 1.42) was perceived to be significantly more uncertain than numeric (*M*_numeric_ = 3.94, *SD* = 1.35; *p* < .001, *d* = .36) or no uncertainty (*M*_control_ = 3.99, *SD* = 1.39; *p* < .001, *d* = .23). There was no significant difference between uncertainty ratings in the control and numeric conditions. Pairwise differences are displayed graphically in Figure 3A (note that for clarity the *p* < .001 pairwise comparisons are collapsed across the epistemic and aleatory conditions).

**Figure 3:**
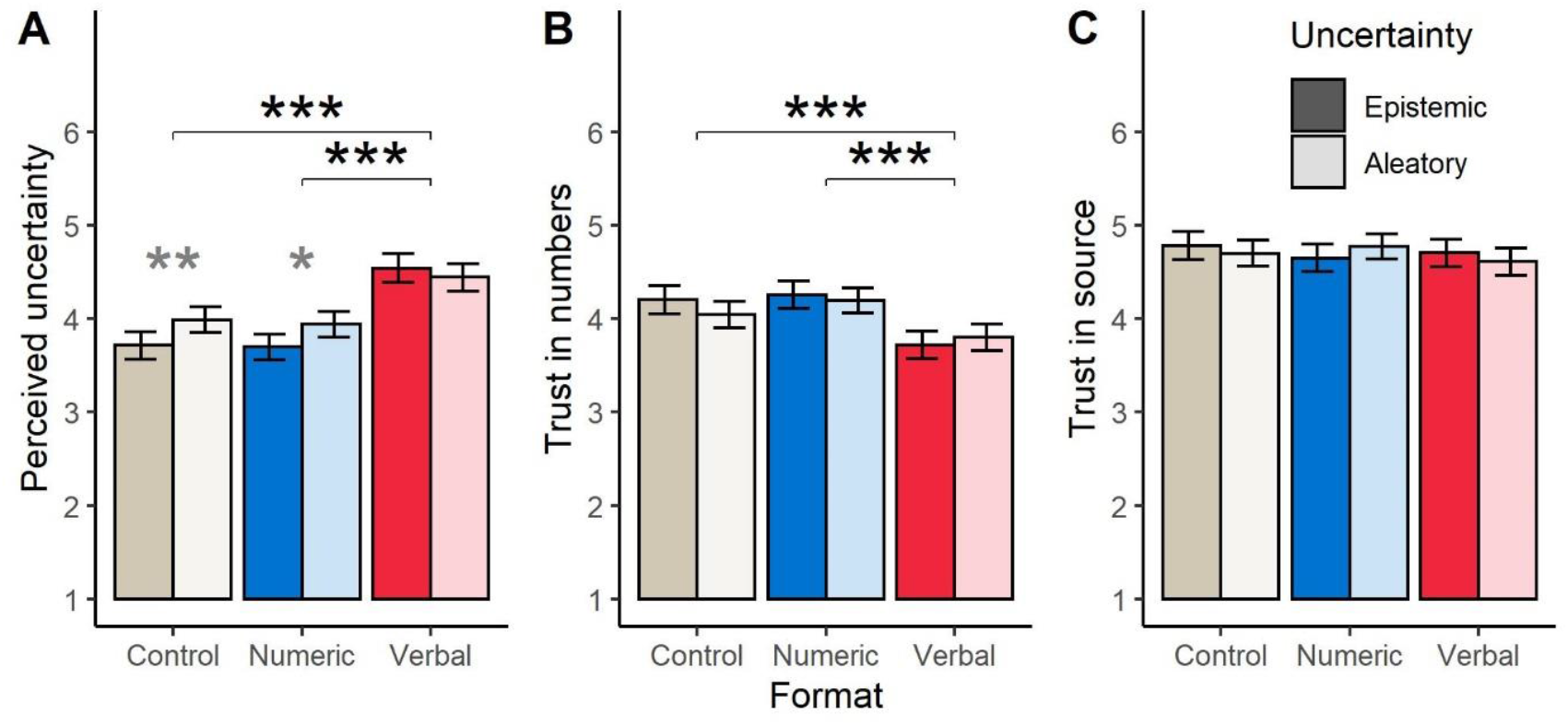
The effect of uncertainty format and type on: (A) perceived uncertainty, (B) trust in numbers, and (C) trust in source (means and 95% CI). Horizontal bars indicate a significant pairwise difference between format conditions (for both epistemic and aleatory uncertainty types), greyed stars indicate significant pairwise difference between epistemic and aleatory uncertainty types within condition, \**p* < .05, \*\**p* < .01, \*\*\**p* < .001.

We also examined pairwise differences between epistemic and aleatory uncertainty types within each of the three condition groups (means reported above). Among participants in the control format condition (who read no additional uncertainty information), aleatory uncertainty was perceived to be more uncertain than epistemic uncertainty (*p* = .008, *d* = .19). Among participants presented with the numeric condition, aleatory uncertainty was also perceived to be more uncertain than epistemic uncertainty (*p* = .014, *d* = .17). In the verbal uncertainty condition, there was no significant difference in perceived uncertainty between aleatory and epistemic uncertainty types.

Considering *trust in the numbers*, a two-way ANOVA found no significant main effect of uncertainty type (i.e. no significant difference between epistemic and aleatory uncertainty across conditions; *F*(1, 2302) = 0.62, *p* = 0.43). We did find a significant main effect of uncertainty format, *F*(2, 2302) = 22.69, *p* < .001, η _*p*_^2^ = .019. There was no significant interaction effect.

Post-hoc tests revealed that participants who read a verbal uncertainty communication trusted the number less compared to participants who read a numeric uncertainty communication (*M*_numeric_ = 4.23, *SD* = 1.38; *M*_verbal_ = 3.76, *SD* = 1.43; *p* < .001, *d* = .33) and a communication with no uncertainty (*M*_control_ = 4.12, *SD* = 1.45; *p* < .001, *d* = .25). There was no significant difference between control and numeric conditions (Figure 3B).

Lastly, considering *trust in the source* of the information, a two-way ANOVA showed no significant effects of uncertainty type (*F*(1, 2299) = 0.08, *p* = 0.77) or of uncertainty format (*F*(2, 2299) = 0.70, *p* = 0.50): participants’ trust in the people responsible for producing the number was not affected by the inclusion of uncertainty in any form (Figure 3C).

Additional analyses focusing on secondary outcomes (difficulty of understanding, emotional response, and perceived competence of source) found no significant effects of either experimental factor (see supplementary information, Appendix 4).

### Interim Discussion

Participants in Study 2 perceived communications including verbal uncertainty as more uncertain than communications without, and were less trusting of the numbers provided, but did not consider the communicators of such information less trustworthy. We also find that the presence or absence of numeric uncertainty has no significant impact on the trust in the number or the source. These effects were consistent across messages communicating either current (epistemic) or future (aleatory) uncertainty about COVID-19 deaths. The results are consistent with our UK findings in Study 1.

Future predictions were perceived as more uncertain than current estimates for control and numeric uncertainty communications, but this framing had no impact on participants’ trust in the numbers or source. We do acknowledge that the aleatory and epistemic stimuli differed in terms of the numbers communicated as we did not want to mislead participants with inaccurate information in the midst of the pandemic (we address this confound in Study 3). As the numeric uncertainty range was substantially larger for the figures in the aleatory condition, we would expect the effect of such a confound to exacerbate, rather than diminish any effects on trust. Thus, we take the current results as evidence that numeric uncertainty, relating to either current or future estimates, does not negatively impact trust in the numbers or the source.

This supports previous work that has demonstrated no negative effect of communicating aleatory uncertainty on trust (Joslyn & LeClerc, 2012; Kuhn, 2000). Although work in this area is not consistent, with some studies finding both positive and negative effects on trust (Johnson & Slovic, 1995), and others showing that the nature of trust effects break down according to participant characteristics such as education level (Schapira et al., 2001).

Considering the between-country differences reported in Study 1 we must also caveat these findings with the limitation that data was only collected from UK residents. We cannot generalise these effects to other cultural contexts where attitudes to uncertainty may differ.

## Study 3

Study 2 examined the impact of both uncertainty type (aleatory, epistemic) and format (numeric, verbal) on trust in COVID-19 information and its source. These two factors have not been examined simultaneously in prior research, to our knowledge. However, as detailed in the introduction, COVID-19 may present an unusual context because of its extreme emotional salience. Do these results extend to other issues, which participants may perceive as more psychologically distant? In this study we present results from an earlier (pre-COVID-19) study investigating the effects of uncertainty type and format in relation to statistics in three other domains, namely climate science, tiger conservation, and unemployment.

The stimuli in this study also avoid the confound in Study 2 arising from the use of differing numbers and ranges for aleatory and epistemic uncertainty.

### Methods

A sample of 2,250 UK adult participants were recruited via the Prolific platform (prolific.co) in June 2019. The study was approved by the University of Cambridge’s Psychology Research Ethics Committee (PRE.2018.041). The sample consisted of 68.6% females and 31.0% males (nine participants (.039%) reported ‘other’ or ‘prefer not to say’) between 18 and 86 years of age (*M* = 36.6, SD = 12.53).

Participants completed an online survey experiment on the Qualtrics platform. Following informed consent, participants were randomly assigned to one of 18 conditions in a 2(uncertainty type)x3(format)x3(topic) between-subject design. Participants were first asked several questions about their current beliefs relating to the topics examined (not reported here). Participants then read a short paragraph about one of the following issues: the rise of the global surface temperature, the decline of the tiger population in India, or rise of the unemployment rate in the UK. All these statements were framed in a negative direction to control for valence effects (i.e., the statistics in presented each condition could broadly be considered ‘bad news’). For instance, although the UK unemployment rate was declining at the time of the research, we stated that it is rising. The uncertainty about these estimates was either absent (control), included as a numerical range (presented as minimum-maximum range), or as a verbal statement that a (unquantified) level uncertainty exists around the estimate. This information was either presented as the present state for 2019 (epistemic condition) or as a prediction for 2025 for the unemployment and tiger numbers or 2050 for the climate numbers (aleatory condition).

For example, participants assigned to the tigers topic and aleatory numeric uncertainty conditions read the following information: “An official report has come out with new information about the number of tigers in India. It shows that the number of tigers could fall to a historic low of 2,226 (minimum 1,945 to maximum 2,491) by 2025.” Participants assigned to the unemployment topic and epistemic verbal uncertainty conditions read: “An official report has come out with new information about the unemployment rate in the United Kingdom. It shows that the UK’s unemployment rate has risen to 3.8% in the first quarter of 2019. The report states that there is some uncertainty around this estimate, it could be somewhat higher or lower.” The texts for all 18 experimental conditions are reported in Appendix 5.

After reading the information, participants were then asked the same questions as in Studies 1 and 2 to ascertain their perception of the uncertainty in the number, their trust in the number (three items; α = .91) and their trust in the people responsible for producing the number. Upon completion of the survey, participants were debriefed with information about the real statistics and sources of these numbers.

### Results

For simplicity of reporting, and because numbers and time frames varied across topics, we report results for each of the three topics separately, examining the effect of uncertainty type and format with 2×3 two-way ANOVAs as in Study 2. For consistency, we report the results of Tukey’s post hoc tests based on one-way ANOVAs examining the effect of format for epistemic and aleatory types separately (shown in Figures 4-6; cell means and SDs reported in Appendix 5). We note instances where these differ from the two-way ANOVA results (i.e. the pattern of significant pairwise comparisons between formats differs across uncertainty types despite no significant interaction between format and type in two-way ANOVAs). For completeness, we also report pairwise comparisons between uncertainty types and topics, at each level of the other experimental factors, in Appendix 6.

**Figure 4:**
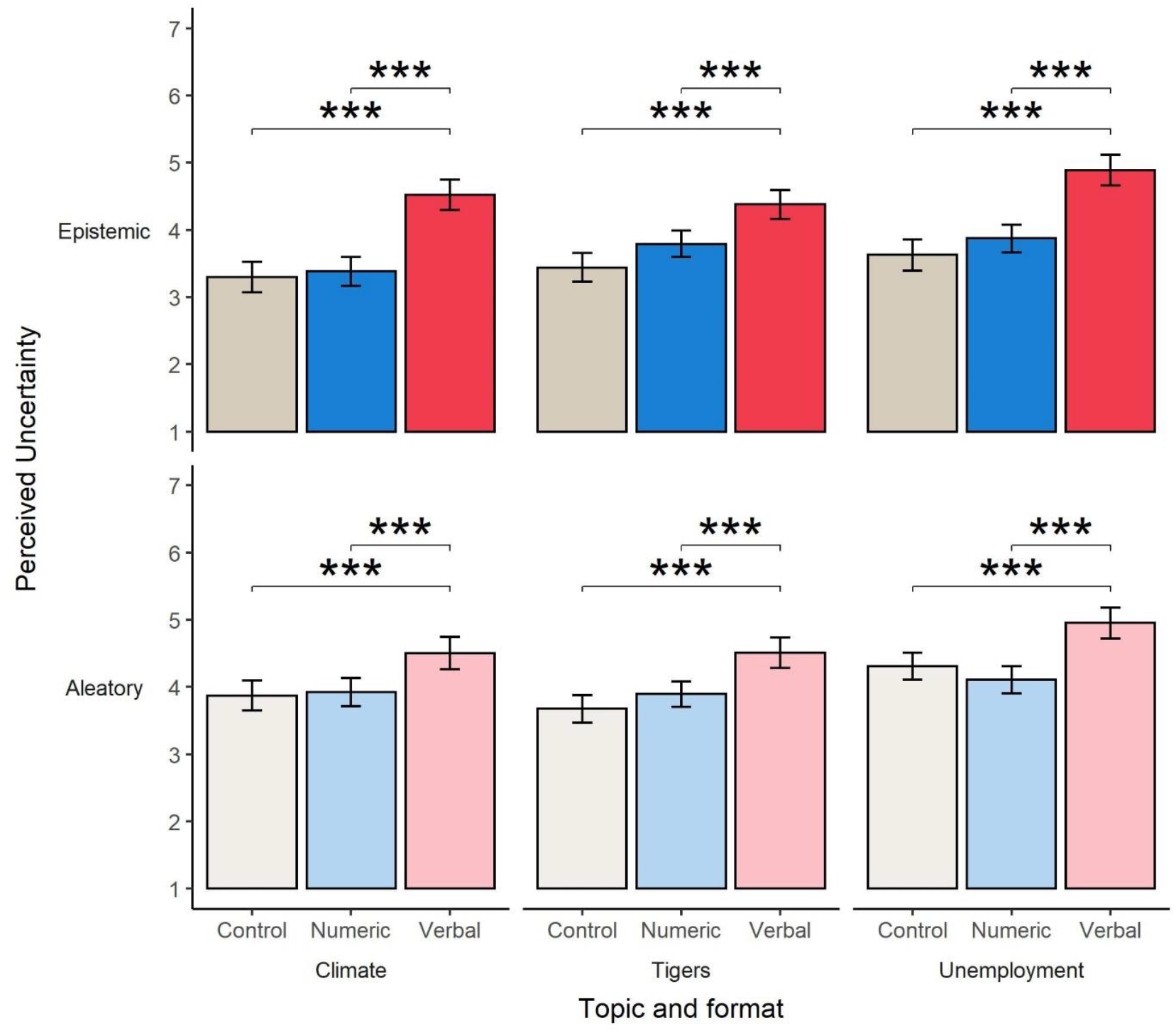
The effect of uncertainty format on perceived uncertainty, plotted across uncertainty type (rows) and topics (columns; means and 95% CI). Horizontal bars indicate a significant pairwise difference between format conditions. \*\*\**p* < .001.

**Figure 5:**
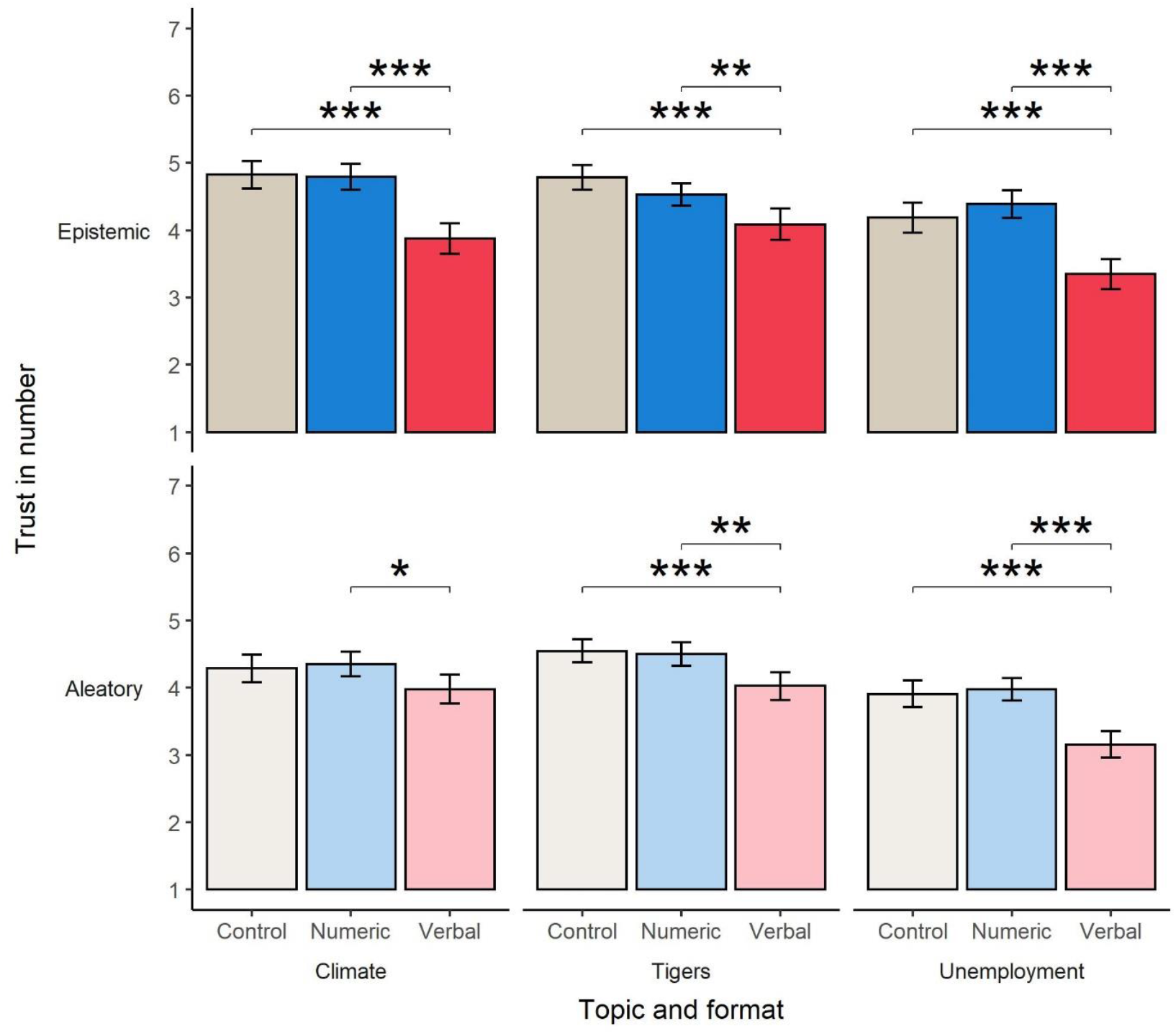
The effect of uncertainty format on trust in numbers, plotted across uncertainty type (rows) and topics (columns; means and 95% CI). Horizontal bars indicate a significant pairwise difference between format conditions. \**p* < .05, \*\**p* < .01\*\*\**p* < .001.

**Figure 6:**
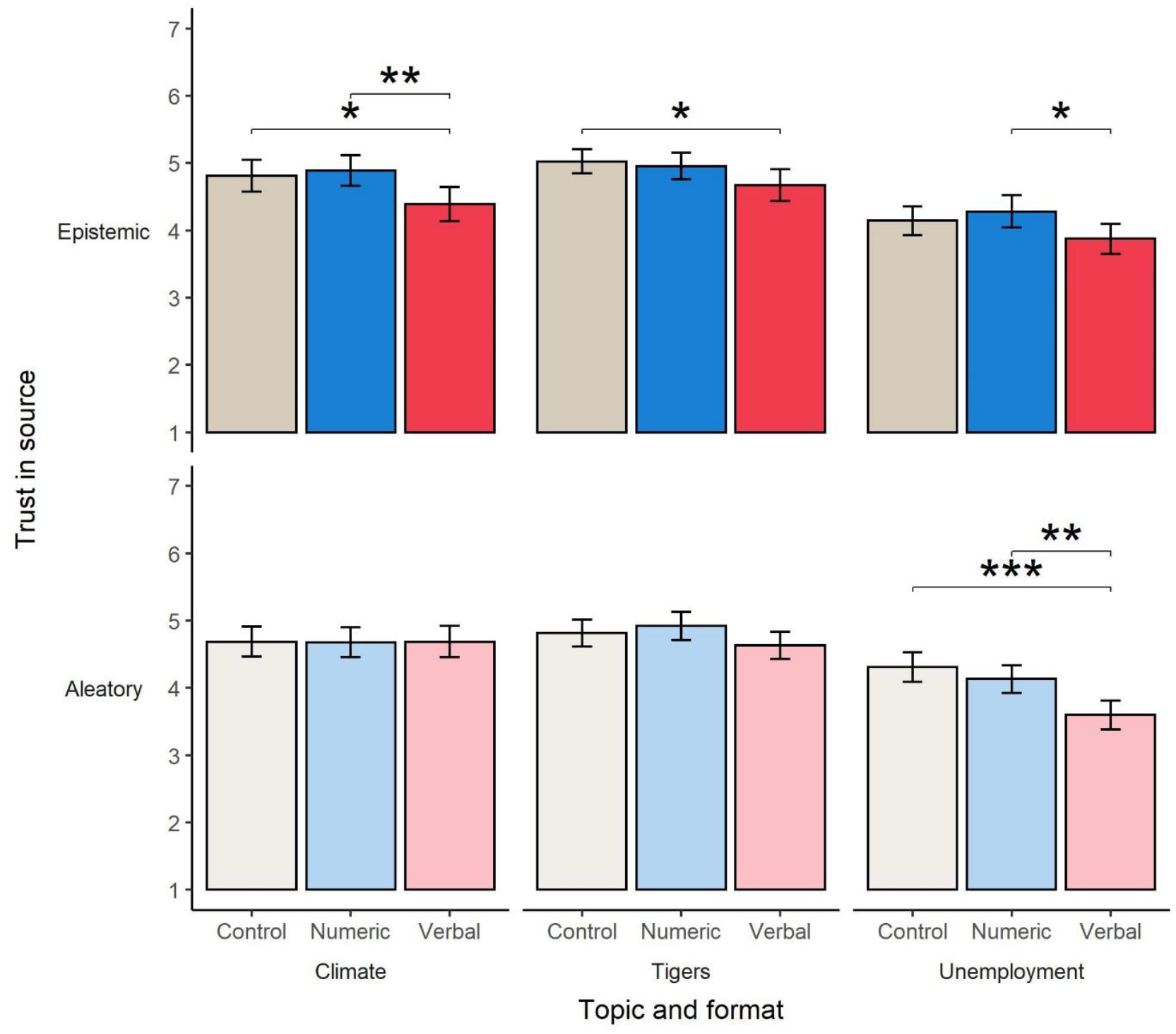
The effect of uncertainty format on trust in source, plotted across uncertainty type (rows) and topics (columns; means and 95% CI). Horizontal bars indicate a significant pairwise difference between format conditions. \**p* < .05, \*\**p* < .01\*\*\**p* < .001.

#### Perceived uncertainty

We consider first the effect of our experimental manipulations on perception of uncertainty. For participants in the climate topic condition, two-way ANOVA results revealed a significant interaction between uncertainty type and format, *F*(2, 747) = 4.41, *p* = .01, η_G_^2^ = 0.012. Separate one-way ANOVAs in the epistemic (*F*(2, 369) = 37.70, *p* < .001, η_G_^2^ = 0.170) and aleatory (*F*(2, 378) = 9.55, *p* < .001, η_G_^2^ = 0.048) uncertainty conditions both revealed a significant effect of format, though this effect was larger in the epistemic condition.

For participants who viewed information about tiger numbers, we report a significant main effect of format, *F*(2, 751) = 36.44, *p* < .001, η_G_^2^ = 0.088, but not uncertainty type, *F*(1, 751) = 3.30, *p* = .07, and no significant interaction.

Among participants reading information about unemployment numbers, we report a significant interaction between uncertainty type and format, *F*(2, 738) = 4.22, *p* = .01, η_G_^2^ = 0.011. Follow-up one-way ANOVAs examining reveal a significant effect of format in the epistemic (*F*(2, 364) = 35.76, *p* < .001, η_G_^2^ = 0.164) and aleatory (*F*(2, 374) = 17.30, *p* < .001, η_G_^2^ = 0.085) uncertainty conditions, with this effect again larger in the epistemic condition.

Across all topics and uncertainty types, post-hoc tests comparing formats revealed a consistent pattern: verbal uncertainty was perceived as more uncertain than the control format (all *p* < .001, *d*s .48-.99) and numeric format (all *p* < .001, *d*s .46-.92), with no significant differences between numeric and control condition.

#### Trust in numbers

Turning to trust in the numbers, we report for participants in the climate condition a significant interaction between uncertainty type and format, *F*(2, 747) = 5.60, *p* < .01, η_G_^2^ = 0.015. Follow up one-way ANOVAs indicated a significant effect of format for both epistemic, *F*(2, 369) = 26.43, *p* < .001, η_G_^2^ = 0.125, and aleatory uncertainty, *F*(2, 378) = 3.80, *p* = .02, η_G_^2^ = 0.020, with a larger effect for epistemic uncertainty. Pairwise comparisons revealed that, for epistemic uncertainty, verbal uncertainty was considered less trustworthy than control (*p* < .001, *d* = .78) or numeric formats (*p* < .001, *d* = .77). For aleatory uncertainty, verbal uncertainty was considered less trustworthy than numeric uncertainty (*p* = .03, *d* = .33; with no other significant differences).

For participants reading information about tiger numbers, we report only a significant main effect of format, *F*(2, 751) = 21.73, *p* < .001, η_G_^2^ = 0.055. In both epistemic and aleatory conditions, post-hoc tests indicate that verbal uncertainty was considered less trustworthy than control (*ps* < .001, *d*_epistemic_ =. 59, *d*_aleatory_ = .48) or numeric uncertainty (*ps* < .01, *d*_epistemic_ = .39, *d*_aleatory_ = .43).

For participants reading information about unemployment numbers, we report a significant main effect of uncertainty type, *F*(1, 738) = 12.51, *p* < .001, η_G_^2^ = 0.017, with aleatory uncertainty considered less trustworthy than epistemic. We also report a significant main effect of format, *F*(2, 738) = 49.20, *p* < .001, η_G_^2^ = 0.118. In both epistemic and aleatory conditions, post-hoc tests indicate that verbal uncertainty was considered less trustworthy than control (*ps* < .001, *d*_epistemic_ =. 68, *d*_aleatory_ = .68) or numeric uncertainty (*ps* < .001, *d*_epistemic_ = .86, *d*_aleatory_ = .78).

#### Trust in source

Lastly, we examine participant’s trust in the source of the information presented in the experiment. Considering participants in the climate topic condition, a 2(type)x3(format) ANOVA revealed no significant main effects but a marginally significant interaction, *F*(2, 747) = 2.68, *p* = .07, η_G_^2^ = 0.007. Follow-up one-way ANOVAs indicated a significant effect of format in the epistemic uncertainty condition, *F*(2, 369) = 5.04, *p* < .01, η_G_^2^ = 0.027, but not aleatory condition, *F*(2, 378) = 0.00, *p* = 1.00. Post hoc tests investigating pairwise differences between formats for epistemic uncertainty revealed that the source of communications including the verbal uncertainty format was less trusted than the source of control (*p* = .03, *d* = .31), or numeric format communications (*p* = .009, *d* =.37).

For participants in the tigers topic condition we report only a significant main effect of format, *F*(2, 751) = 4.84, *p* < .01, η_G_^2^ = 0.013, however separate sets of post hoc tests revealed a significant difference only in the epistemic uncertainty type condition: the source of verbal uncertainty was less trusted than the source of the control communication without uncertainty (*p* = .04, *d* = .30).

Considering participants who read information about unemployment numbers, we report a main effect of format only, *F*(2, 738) = 12.78, *p* < .001, η_G_^2^ = 0.033. However, separate post hoc tests for epistemic and aleatory uncertainty types indicated differing patterns of significant pairwise differences. In the epistemic uncertainty condition, the source of verbal uncertainty was less trusted than source of numeric uncertainty (*p* = .03, *d* = .32). In the aleatory uncertainty condition, the source of verbal uncertainty was less trusted than source of numeric uncertainty (*p* <.001, *d* = .58), and the source of control information without uncertainty (*p* = .001, *d* = .44).

Results for additional secondary outcomes—reported difficulty and emotional response—are reported in Appendix 7.

### Interim discussion

In Study 3 we examined the impact of uncertainty format (a specific, numeric quantification of uncertainty or a vague and unquantified verbal expression of uncertainty) and how this is affected by whether the uncertainty is regarding the current state of affairs (epistemic) or looking to the future (aleatory). We find across all topics, and for both epistemic and aleatory uncertainty, that messages with no uncertainty information and messages with numeric uncertainty (a confidence interval) are perceived as essentially equally uncertain. Expressions of verbal uncertainty, however, are consistently perceived as more uncertain.

Similarly, when considering trust in the numbers presented, we find a consistent pattern across topics and uncertainty types: statistics with no uncertainty or numeric uncertainty do not significantly differ in terms of how much participants trust the numbers, while messages with verbal uncertainty are considered less trustworthy. The one exception to this is future-focused climate predictions, where we report no significant difference between messages without uncertainty and messages with verbal uncertainty, in terms of trust.

Lastly, we find a slightly more heterogenous set of results when examining trust in the (unnamed) source of the numbers. We again report no significant differences between messages with no uncertainty or numeric uncertainty. However, the effect of verbal uncertainty (vs control) differs depending on both the topic and type of uncertainty. When the uncertainty is relating to current climate or tiger statistics, we find that verbal uncertainty leads to lower levels of trust in the source, but there is no effect when uncertainty relates to current employment rate estimates. We find the opposite pattern when considering aleatory uncertainty: verbal uncertainty has no impact on trust in the source of numbers when relating to climate or tiger statistics but does lead to lower trust when numbers relate to unemployment statistics. This suggests that the impact of verbal uncertainty on trust in the information source is context-specific, and caution should be exercised in generalizing effects from one domain to another.

## Discussion

Across three different studies, covering multiple countries, topics, and temporal framings, we find a clear pattern in which communications expressing statistical uncertainty as a range around a point estimate are not perceived less trustworthy nor are the communicators considered less trustworthy. Only when drawing on the statistical power of a combined sample of more than 10,000 participants (Study 1) do we find a small negative effect of such uncertainty on trust in the numbers presented, and still no significant effect on trust in the source. In this regard, our results are consistent with the prior work of van der Bles (2020) and others (see Gustafson & Rice, 2020), and extend these findings to the new, and arguably more personally salient, context of the COVID-19 pandemic. As a further extension on van der Bles (2020), we also find minimal impact of numeric uncertainty on trust when claims relate to future predictions rather than current estimates. First and foremost, then, we echo previous recommendations that communicators should feel confident that they can use numerical ranges around their point estimates when communicating COVID-19 statistics or projections without risking a significant undermining of audience trust (Blastland et al., 2020; van der Bles et al., 2019).

Figures communicated with imprecise, verbal expressions of the existence of uncertainty regarding a statistic (e.g., ‘could be higher or lower’) may be considered less trustworthy and undermine the credibility of communicators. On this front, our results paint a more complicated picture. Regarding *trust in numbers*, the negative impact of this type of uncertainty when applied to COVID-19 statistics varies across countries (Study 1). In the UK we find that such expressions consistently lower trust across types of uncertainty (epistemic or aleatory; study 2) and topics (study 2 and 3). The extent to which this translates into decreased *trust in the source* of the information appears to vary by domain as well as type of uncertainty. Verbal uncertainty had little impact on trust in the source of COVID statistics (Studies 1 & 2) and for other topics effects vary by both type and topic (Study 3). The sources of unquantified verbal uncertainty may be considered trustworthy for some forecast statistics (COVID-19 deaths, climate warming, tiger numbers) but not others (unemployment). A possible explanation for these differences may be that some contexts are viewed as inherently more uncertain, and blunt admissions of uncertainty are considered more acceptable. But taken as whole, these findings suggest that communicators should be cautious in mentioning of the existence of uncertainty regarding a statistic without some description of the bounds of that uncertainty.

In no case did we find that expressions of uncertainty *increase* trust in numbers or their source. This is noteworthy as some previous research has found provision of uncertainty to increase credibility in some cases, for example in earthquake and climate projections (Joslyn & Demnitz, 2019; Joslyn & LeClerc, 2016; Nakayachi et al., 2018). However other studies have found no effect of uncertainty on credibility in other domains (e.g. GM food, Gustafson & Rice, 2019; health, Lipkus et al., 2001). These mixed findings are not surprising given the range of issues, forms of uncertainty and operationalizations of trust and credibility used in previous research (Gustafson & Rice, 2020). The current study contributes to ongoing work in identifying the broader factors that determine when communication of uncertainty does or does not garner greater trust.

While a single message with uncertainty may not increase trust, it might buffer against future damage to credibility if figures are revised. Batteux et al. (2021) report that the inclusion of a range (vs point estimate) in COVID-19 vaccine efficacy communications had no immediate impact on trust. However, when participants were later presented with conflicting evidence (updating the previous estimate), those who had first received a point estimate with no uncertainty reported lower trust in the communicator, compared to those receiving a range. This suggests that communicating uncertainty up front can mitigate later skepticism when scientific evidence is updated. Such findings are particularly relevant to situations such as the COVID-19 pandemic, where evidence and recommendations are constantly updated (Kreps & Kriner, 2020; Martin et al., 2020).

The consistency of our findings regarding numeric uncertainty and trust—across countries, issues, and temporal framings—gives us some confidence in our conclusions. However, we must acknowledge some limitations. Firstly, the uncertainty ranges communicated in our messages were all drawn from existing reports. While this provides some ecological validity, it did mean we were not able to control for the magnitude of the range presented across some conditions. Previous research suggests people may be less trusting of uncertainty presented as a very large numeric range (Kreps & Kriner, 2020; van der Bles et al., 2020). More research is needed to identify the effect of range magnitude and how this varies across contexts.

Secondly, Studies 2 and 3 were only conducted in UK samples. As shown in Study 1, there is between-country variation in how people respond to uncertainty. Therefore, we cannot confidently generalize our UK findings to other national contexts. Future research should seek to replicate these findings with other countries as well as investigating possible explanations for the variation seen in Study 1.

Lastly, the types of uncertainty we examined in the current research were narrowly defined. Thus, our findings may not map onto to other forms or formats of uncertainty. For example, statistical uncertainty could be expressed visually—rather than as numerals—and the existence of unquantified uncertainty could be phrased in a multitude of different ways. Uncertainty can also be further attributed to different sources, with potentially differing impacts. For example, claims could be accompanied by statements about uncertainty due to a lack of data, poor quality research, disagreement among experts, or the nature of the scientific method itself. The inclusion of such explanations may moderate the effects of uncertainty on credibility and deserves further attention.

In conclusion, we find that acknowledging uncertainty in the form a range has minimal impact on the perceived trustworthiness of figures or their assumed source. This applies to COVID-19 statistics and claims in other domains, regardless of whether the figures relate to current estimates or future projections. Statements merely acknowledging the existence of uncertainty may be treated with more skepticism, though this appears to be context dependent.

## Data Availability

Data and R analysis scripts are available on the Open Science Foundation website.

https://osf.io/y982k/?view_only=07f2c4fd8000488f90f564f5a209bb9c

## Data availability

Data and R analysis scripts are available on the Open Science Foundation website: https://osf.io/y982k/?view_only=07f2c4fd8000488f90f564f5a209bb9c

## Acknowledgements

We would like to thank María del Carmen Climént Palmer, Ban Mutsuhisa, Jin Park, and Giulia Luoni for additional translations, the University of Tokyo for their collaboration, and all the participants and those who helped to administer the study.

## Footnotes

1 We acknowledge that the inclusion of the qualifier “about” in the sentence presented introduces some level of uncertainty to all participants. However we note this is consistent across conditions and previous research (van der Bles et al., 2020) has reported sentences with such qualifiers are not perceived as more uncertain or less trusted than statements without these words.

2 Given that the sample collected in France was much larger than all other samples and would have had more influence on the combined results, we repeated these analyses with the French sample excluded. The pattern of significant results remained the same.

