## Supplementary information for "The effects of communicating uncertainty around statistics on public trust: an international study"

**Appendix 1: Study 1 secondary outcomes**

We also asked participants in Study 1 two additional questions:

- How does the information you just saw make you feel? (0 = Negative/unhappy, 10 = positive/happy)
- How easy or difficult do you find this information to understand? (1 = Very difficult, 7 = Very easy)

A one-way ANOVA conducted on the combined sample indicated that *reported feelings* differed across experimental conditions, $F(2, 10489) = 4.99$, $p < .01$, $\eta^2 = 0.001$. Post hoc analyses revealed that participants in the verbal uncertainty condition reported that the information made them feel, on average, more positive ($M_{\text{verbal}} = 4.95$, $SD = 2.39$) than those in the numeric uncertainty ($M_{\text{numeric}} = 4.79$, $SD = 2.40$; $M_{\text{diff}} = 0.16$, 95CI [0.02, 0.29], $p < .05$, $d = -.07$) and control conditions ($M_{\text{control}} = 4.79$, $SD = 2.51$; $M_{\text{diff}} = 0.16$, 95CI [0.02, 0.30], $p < .05$, $d = .07$).

Reported *difficulty of understanding* the information also differed across conditions, $F(2, 10472) = 60.87$, $p < .001$, $\eta^2 = 0.011$. Post hoc analyses indicated that all conditions differed significantly from each other; participants in the control condition rated the information easiest to understand ($M_{\text{control}} = 5.20$, $SD = 1.46$), followed by those in the verbal ($M_{\text{verbal}} = 4.96$, $SD = 1.46$), then numeric uncertainty conditions ($M_{\text{numeric}} = 4.82$, $SD = 1.44$; $M_{\text{diff:control-numeric}} = -0.38$, 95CI [-0.46, -0.3], $p < .001$, $d = .26$; $M_{\text{diff:control-verbal}} = -0.24$, 95CI [-0.32, -0.16], $p < .001$, $d = .16$; $M_{\text{diff:numeric-verbal}} = 0.14$, 95CI [0.06, 0.22], $p < .001$, $d = .10$).

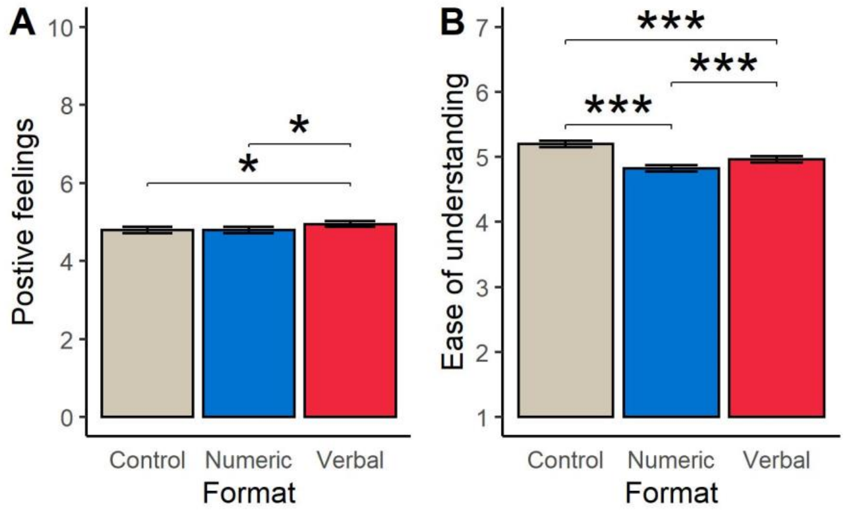

Figure S1: The effect of experimental condition on (A) reported feelings in response to the information and (B) rating of difficulty of understanding. Means and 95%Cis shown. Horizontal bars indicate a significant pairwise differences between conditions, **p* < .05, ****p* < .001

**Appendix 2: Study 1 ANOVA results by country**

Table S1: Results of ANOVA analyses of the effect of experimental condition on Study 1 primary outcomes, for each country.

| Outcome | Sample | *Df* | *F* | *p* | $\eta^2$ |
|---|---|---|---|---|---|
| Perceived uncertainty | Australia | 2, 665 | 2.60 | 0.08 | 0.008 |
| | China | 2, 692 | 0.41 | 0.67 | 0.001 |
| | France | 2, 2999 | 6.36 | < .01 | 0.004 |
| | Germany | 2, 681 | 7.01 | < .001 | 0.02 |
| | Italy | 2, 602 | 8.18 | < .001 | 0.026 |
| | Japan | 2, 694 | 2.73 | 0.07 | 0.008 |
| | S. Korea | 2, 696 | 4.09 | 0.02 | 0.012 |
| | Mexico | 2, 650 | 2.55 | 0.08 | 0.008 |
| | Spain | 2, 684 | 9.54 | < .001 | 0.027 |
| | Sweden | 2, 678 | 1.66 | 0.19 | 0.005 |
| | UK | 2, 700 | 23.09 | < .001 | 0.062 |
| | US | 2, 698 | 28.27 | < .001 | 0.075 |
| Trust in numbers | Australia | 2, 664 | 2.94 | 0.05 | 0.009 |
| | China | 2, 694 | 4.52 | 0.01 | 0.013 |
| | France | 2, 2999 | 4.45 | 0.01 | 0.003 |
| | Germany | 2, 678 | 3.50 | 0.03 | 0.010 |
| | Italy | 2, 597 | 1.17 | 0.31 | 0.004 |
| | Japan | 2, 691 | 2.87 | 0.06 | 0.008 |
| | S. Korea | 2, 696 | 11.99 | < .001 | 0.033 |
| | Mexico | 2, 648 | 2.03 | 0.13 | 0.006 |
| | Spain | 2, 681 | 2.33 | 0.10 | 0.007 |
| | Sweden | 2, 676 | 2.86 | 0.06 | 0.008 |
| | UK | 2, 700 | 14.05 | < .001 | 0.039 |
| | US | 2, 697 | 5.87 | < .01 | 0.017 |
| Trust in source | Australia | 2, 663 | 2.24 | 0.11 | 0.007 |
| | China | 2, 694 | 0.72 | 0.49 | 0.002 |
| | France | 2, 2999 | 0.41 | 0.67 | 0.000 |
| | Germany | 2, 682 | 2.59 | 0.08 | 0.008 |
| | Italy | 2, 594 | 0.76 | 0.47 | 0.003 |
| | Japan | 2, 694 | 1.00 | 0.37 | 0.003 |
| | S. Korea | 2, 697 | 1.89 | 0.15 | 0.005 |
| | Mexico | 2, 652 | 0.66 | 0.52 | 0.002 |
| | Spain | 2, 685 | 0.70 | 0.50 | 0.002 |
| | Sweden | 2, 677 | 1.62 | 0.20 | 0.005 |
| | UK | 2, 699 | 1.79 | 0.17 | 0.005 |
| | US | 2, 698 | 1.79 | 0.17 | 0.005 |

**Appendix 3: Study 1 exploratory analyses**

There was considerable variation in the magnitude of experimental effects between country samples. To explore potential explanations for this variation we examined the correlation between effect sizes (as Cohen's *d*) and several different country-level variables drawn from different sources.

Uncertainty Avoidance Index: We drew on the most recent Uncertainty Avoidance values provided by Hofstede's website (scaled 0-100; dated 2015). According Hofstede Insights (https://hi.hofstede-insights.com/national-culture), "The Uncertainty Avoidance dimension expresses the degree to which the members of a society feel uncomfortable with uncertainty and ambiguity".

Risk attitude: We also drew on data from the World Risk Poll (https://wrp.lrfoundation.org.uk) to capture broad measures of country-level attitude towards risk and uncertainty. Participants in this global survey were asked the following question, "When you hear the word RISK, do you think more about opportunity or danger?", with responses coded as: opportunity, danger, both, neither, don't know or refused. Using provided weights, we calculated the population percentages responding either 'opportunity' or 'danger' as separate country-level variables.

Numeracy: Participants completing the Study 1 survey experiment also competed a 1-5 measure of numeracy, comprised of the Adaptive Berlin Numeracy Test (scored 1-4; Cokely et al., 2012) and additional numeracy item drawn from Lipkus et al (2001; 'Which represents the greatest risk?' 1 in 100, 1 in 10, 1 in 1000; scored 0-1). The mean numeracy score for each country sample was calculated.

General Social Trust: Study 1 participants also completed a measure of General Social Trust taken from the General Social Survey: 'Generally speaking, would you say most people can be trusted, or that you can't be too careful in dealing with people?' Responses were collected with a 7-point scale (1= You can't be too careful in dealing with people, 7 = Most people can be trusted). The mean trust score for each country sample was calculated.

Table S2 reports the experimental effects sizes across countries for each outcome of interest and contrast (numeric or verbal vs. control) alongside collected country-level variables.

To examine the extent to which these country/sample-level variables explain the inter-country variation in effect sizes, we calculated the correlation between effect size and explanatory variable for each outcome. Given these analyses are based only on twelve datapoints, we present these results for exploratory purposes only. As seen in table S3, several moderately sized correlations were detected, however only one was significant at the $p < 0.5$ level. There was a significant negative correlation between the effect of a verbal message (vs control) on trust in numbers and sample mean level of numeracy; country samples which were, on average, more numerate rated statistics including verbal expression of uncertainty as less trustworthy (relative to control), compared to less numerate samples ($r = -.71$, $p = .009$). We illustrate this relationship in Figure S2.

Table S2: Effect sizes by country with investigated explanatory variables

| Country | Effect size (outcome and condition vs. control) | | | | | | UAI | Risk = opportunity (%)[a] | Risk = danger (%)[a] | GST | Numeracy |
|---|---|---|---|---|---|---|---|---|---|---|---|
| | Perceived uncertainty | | Trust in numbers | | Trust in source | | | | | | |
| | Numeric | Verbal | Numeric | Verbal | Numeric | Verbal | | | | | |
| Australia | 0.20 | 0.27 | -0.11 | -0.29 | -0.15 | -0.24 | 51 | 22.40 | 74.08 | 3.96 | 2.41 |
| China | 0.05 | 0.12 | -0.21 | -0.36 | -0.06 | -0.14 | 30 | 20.77 | 43.62 | 4.95 | 2.83 |
| France | 0.05 | 0.2 | -0.08 | -0.18 | -0.02 | -0.06 | 86 | 19.15 | 77.26 | 2.98 | 2.44 |
| Germany | 0.22 | 0.48 | -0.25 | -0.3 | -0.24 | -0.20 | 65 | 39.18 | 55.38 | 3.62 | 2.53 |
| Italy | 0.53 | 0.45 | -0.17 | -0.16 | -0.15 | -0.03 | 75 | 17.58 | 77.09 | 3.65 | 2.13 |
| Japan | -0.14 | 0.11 | 0.09 | -0.16 | -0.05 | -0.16 | 92 | 14.50 | 75.34 | 3.84 | 2.77 |
| S. Korea | 0.08 | 0.31 | -0.51 | -0.18 | 0.05 | -0.15 | 85 | 36.69 | 57.62 | 3.97 | 2.60 |
| Mexico | 0.09 | 0.3 | -0.19 | -0.27 | -0.08 | -0.17 | 82 | 10.12 | 88.25 | 2.82 | 2.16 |
| Spain | 0.31 | 0.53 | -0.23 | -0.24 | -0.07 | -0.14 | 86 | 11.99 | 86.32 | 3.47 | 2.39 |
| Sweden | -0.08 | 0.15 | 0.18 | -0.12 | 0.20 | 0.01 | 29 | 16.66 | 80.98 | 3.73 | 2.52 |
| UK | 0.33 | 0.77 | -0.21 | -0.59 | -0.14 | -0.22 | 35 | 28.21 | 63.78 | 4.04 | 3.22 |
| US | 0.57 | 0.94 | -0.20 | -0.43 | -0.12 | -0.22 | 46 | 34.16 | 63.85 | 4.01 | 3.14 |

UAI = Uncertainty Avoidance Index, GST = General Social Trust.

[a]Estimated percentage of country population selecting response for risk attitudes item (World Risk Poll).

Table S3: Correlations between effect size and explanatory variables

| Outcome | Effect (Condition vs. control) | UAI | | Risk = opportunity | | Risk = danger | | GST | | Numeracy | |
|---|---|---|---|---|---|---|---|---|---|---|---|
| | | *r* | *p* | *r* | *p* | *r* | *p* | *r* | *p* | *r* | *p* |
| Perceived uncertainty | Numeric | -0.12 | 0.72 | 0.31 | 0.32 | -0.06 | 0.86 | 0.04 | 0.89 | 0.12 | 0.70 |
| | Verbal | -0.20 | 0.54 | 0.46 | 0.13 | -0.10 | 0.75 | 0.03 | 0.93 | 0.48 | 0.11 |
| Trust in numbers | Numeric | -0.21 | 0.51 | -0.55 | 0.07 | 0.45 | 0.14 | -0.12 | 0.70 | -0.05 | 0.88 |
| | Verbal | 0.53 | 0.08 | -0.39 | 0.21 | 0.43 | 0.16 | -0.37 | 0.23 | -0.71 | 0.01 |
| Trust in source | Numeric | -0.10 | 0.77 | -0.29 | 0.36 | 0.23 | 0.46 | -0.03 | 0.94 | -0.06 | 0.86 |
| | Verbal | 0.10 | 0.77 | -0.45 | 0.15 | 0.35 | 0.27 | -0.25 | 0.44 | -0.48 | 0.12 |

UAI = Uncertainty Avoidance Index, GST = General Social Trust.

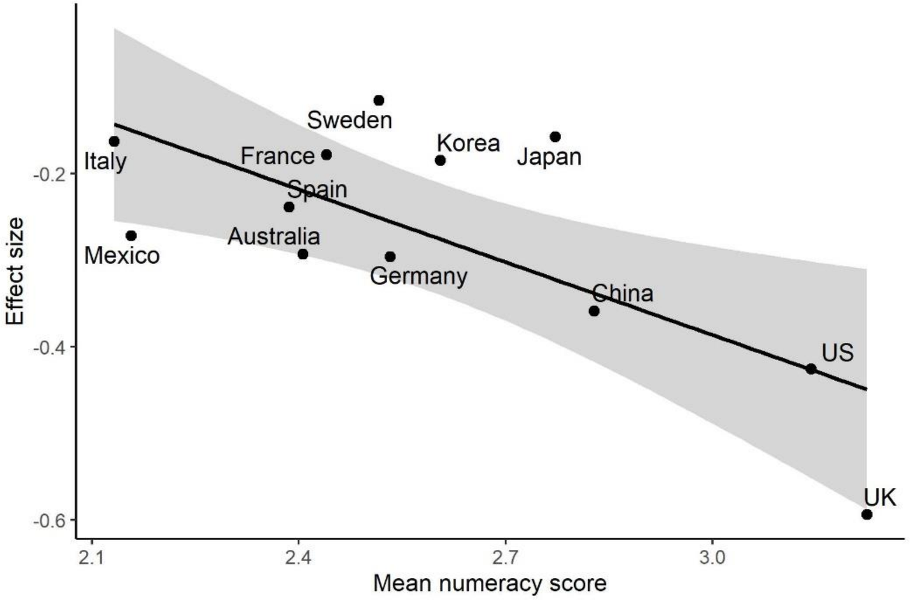

Figure S2. Correlation between effect size of verbal condition (vs. control) on trust in numbers and mean sample numeracy.

**Appendix 4: Study 2 secondary outcomes**

We asked participants in Study 2 several additional questions:

- How does the information you just saw make you feel? (0 = Negative/unhappy, 10 = positive/happy)
- How easy or difficult do you find this information to understand? (1 = Very difficult, 7 = Very easy)
- To what extent do you think that the people responsible for producing this number are competent? (1 = Not at all competent, 7 = Very competent)

Two-way ANOVAs indicated that neither uncertainty condition (control, numeric, verbal) nor uncertainty type (epistemic, aleatory) had a significant main effect on how the information made them feel, ratings of how difficult the information was to understand or perceived competence of the source of the information. There were no significant interactions (all *F*s(1-2, 2301-2302) 0.03-1.14, *p*s > .32; see Figure S3).

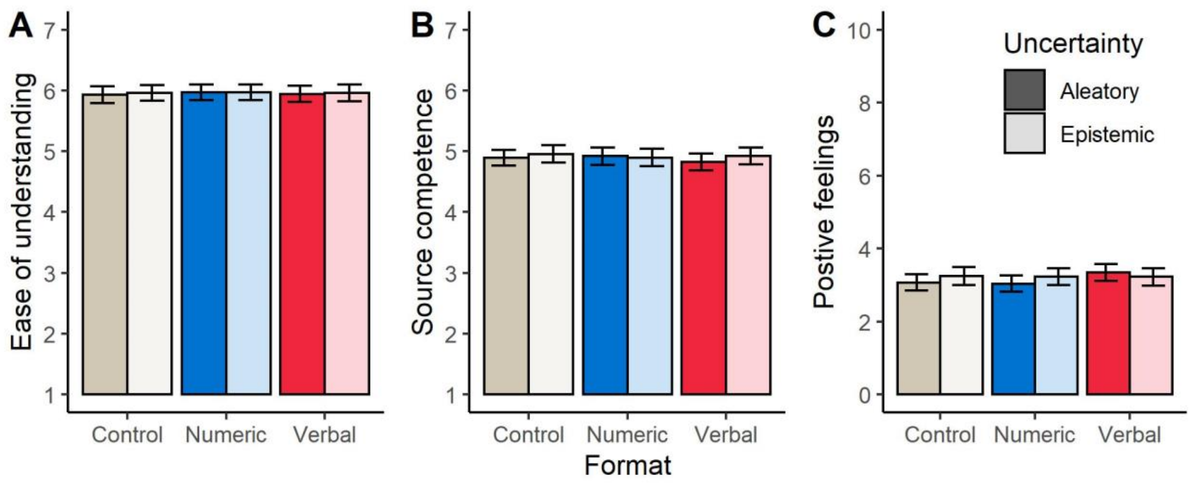

Figure S3: Effect of experimental condition and uncertainty type on: (A) ease of understanding, (B) perceived competence of information source, (C) and reported feelings after reading the information. Means and 95% CI shown.

**Appendix 5: Study 3 materials and results**

Table S4. Study 3 experimental stimuli.

| Topic | Uncertainty type | Uncertainty format | | |
|---|---|---|---|---|
| | | Control | Numeric | Verbal |
| **Climate:** An official report has come out with new information about global warming. It shows that the average surface temperature of the earth… | Epistemic | …has risen 0.87°C in the past 50 years. | … has risen 0.87°C (minimum 0.75 to maximum 0.99°C) in the past 50 years. | …has risen 0.87°C in the past 50 years. The report states that there is some uncertainty around this estimate, it could be somewhat higher or lower. |
| | Aleatory | …is set to rise by 0.87°C in the next 50 years. | …is set to rise by 0.87°C (minimum 0.75 to maximum 0.99°C) in the next 50 years. | …is set to rise by 0.87°C in the next 50 years. The report states that there is some uncertainty around this estimate, it could be somewhat higher or lower. |
| **Tigers:** An official report has come out with new information about the number of tigers in India. It shows that the number of tigers… | Epistemic | …has fallen to a historic low of 2,226 in 2019 | …has fallen to a historic low of 2,226 (minimum 1,945 to maximum 2,491) in 2019. | …has fallen to a historic low of 2,226 in 2019. The report states that there is some uncertainty around this estimate, it could be somewhat higher or lower. |
| | Aleatory | …could fall to a historic low of 2,226 by 2025. | …could fall to a historic low of 2,226 (minimum 1,945 to maximum 2,491) by 2025. | …could fall to a historic low of 2,226 by 2025. The report states that there is some uncertainty around this estimate, it could be somewhat higher or lower. |
| **Unemployment:** An official report has come out with new information about the unemployment rate in the United Kingdom. It shows that the UK's unemployment rate… | Epistemic | …has risen to 3.8% in the first quarter of 2019. | …has risen to 3.8% (minimum 3.6% to maximum 4.0%) in the first quarter of 2019. | …has risen to 3.8% in the first quarter of 2019. The report states that there is some uncertainty around this estimate, it could be somewhat higher or lower. |
| | Aleatory | …could rise to 3.8% by the end of 2025. | …could rise as high as 3.8% (minimum 3.6% to maximum 4.0%) by the end of 2025. | …could rise as high as 3.8% by the end of 2025. The report states that there is some uncertainty around this estimate, it could be somewhat higher or lower. |

Note: Each of the 18 experimental messages consisted of the 'topic' stem followed by the text specified by 'type' row and 'format' column..

Table S5. Study 3 primary outcomes across experimental conditions (mean (SD))

| Conditions | | | n | Perceived uncertainty | Trust in number | Trust in source |
|---|---|---|---|---|---|---|
| Topic | Type | Format | | | | |
| Climate | Epistemic | Control | 121 | 3.30 (1.25) | 4.83 (1.15) | 4.81 (1.29) |
| | | Numeric | 125 | 3.38 (1.20) | 4.80 (1.09) | 4.89 (1.28) |
| | | Verbal | 126 | 4.52 (1.28) | 3.88 (1.27) | 4.39 (1.43) |
| | Aleatory | Control | 128 | 3.88 (1.27) | 4.29 (1.17) | 4.69 (1.28) |
| | | Numeric | 128 | 3.92 (1.19) | 4.35 (1.05) | 4.68 (1.29) |
| | | Verbal | 125 | 4.50 (1.36) | 3.98 (1.21) | 4.69 (1.30) |
| Tigers | Epistemic | Control | 127 | 3.44 (1.21) | 4.78 (1.04) | 5.02 (1.03) |
| | | Numeric | 125 | 3.79 (1.12) | 4.53 (0.95) | 4.95 (1.11) |
| | | Verbal | 124 | 4.38 (1.22) | 4.09 (1.28) | 4.67 (1.33) |
| | Aleatory | Control | 127 | 3.68 (1.18) | 4.55 (0.99) | 4.82 (1.14) |
| | | Numeric | 126 | 3.90 (1.07) | 4.50 (1.00) | 4.92 (1.19) |
| | | Verbal | 128 | 4.51 (1.30) | 4.03 (1.18) | 4.63 (1.14) |
| Unemployment | Epistemic | Control | 118 | 3.63 (1.28) | 4.19 (1.20) | 4.14 (1.19) |
| | | Numeric | 125 | 3.87 (1.15) | 4.39 (1.16) | 4.28 (1.34) |
| | | Verbal | 124 | 4.89 (1.28) | 3.35 (1.25) | 3.87 (1.25) |
| | Aleatory | Control | 122 | 4.31 (1.12) | 3.91 (1.09) | 4.31 (1.23) |
| | | Numeric | 128 | 4.11 (1.13) | 3.97 (0.95) | 4.13 (1.18) |
| | | Verbal | 127 | 4.95 (1.31) | 3.16 (1.12) | 3.60 (1.22) |

**Appendix 6: Study 3 alternative pairwise comparisons**

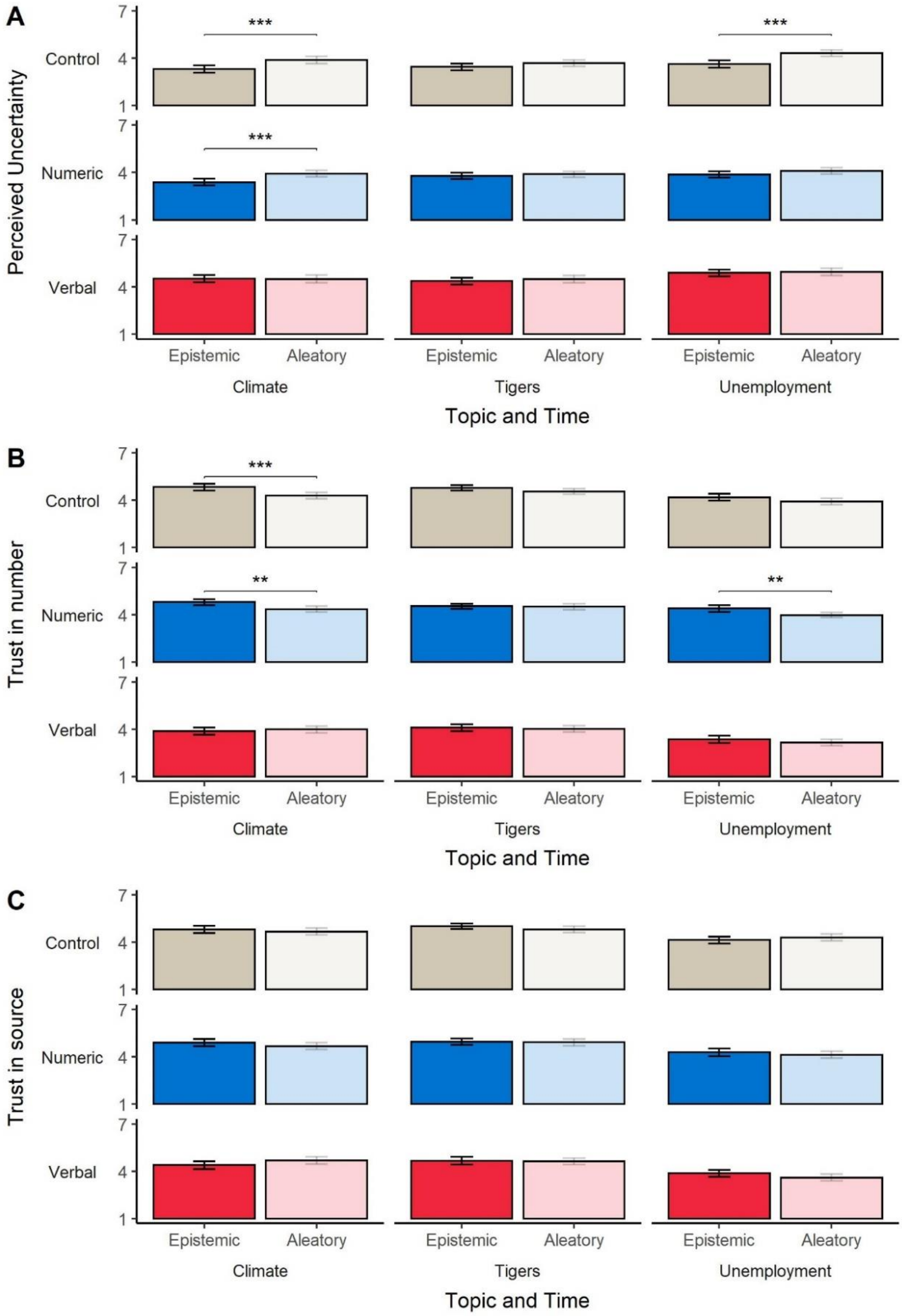

Figure S4: The effect of uncertainty type on: (A) perceived uncertainty, (B) trust in numbers, and (C) trust in source (means and 95% CI). Horizontal bars indicate a significant pairwise difference between epistemic and aleatory conditions (unadjusted). $^{**}p < .01$, $^{***}p < .001$.

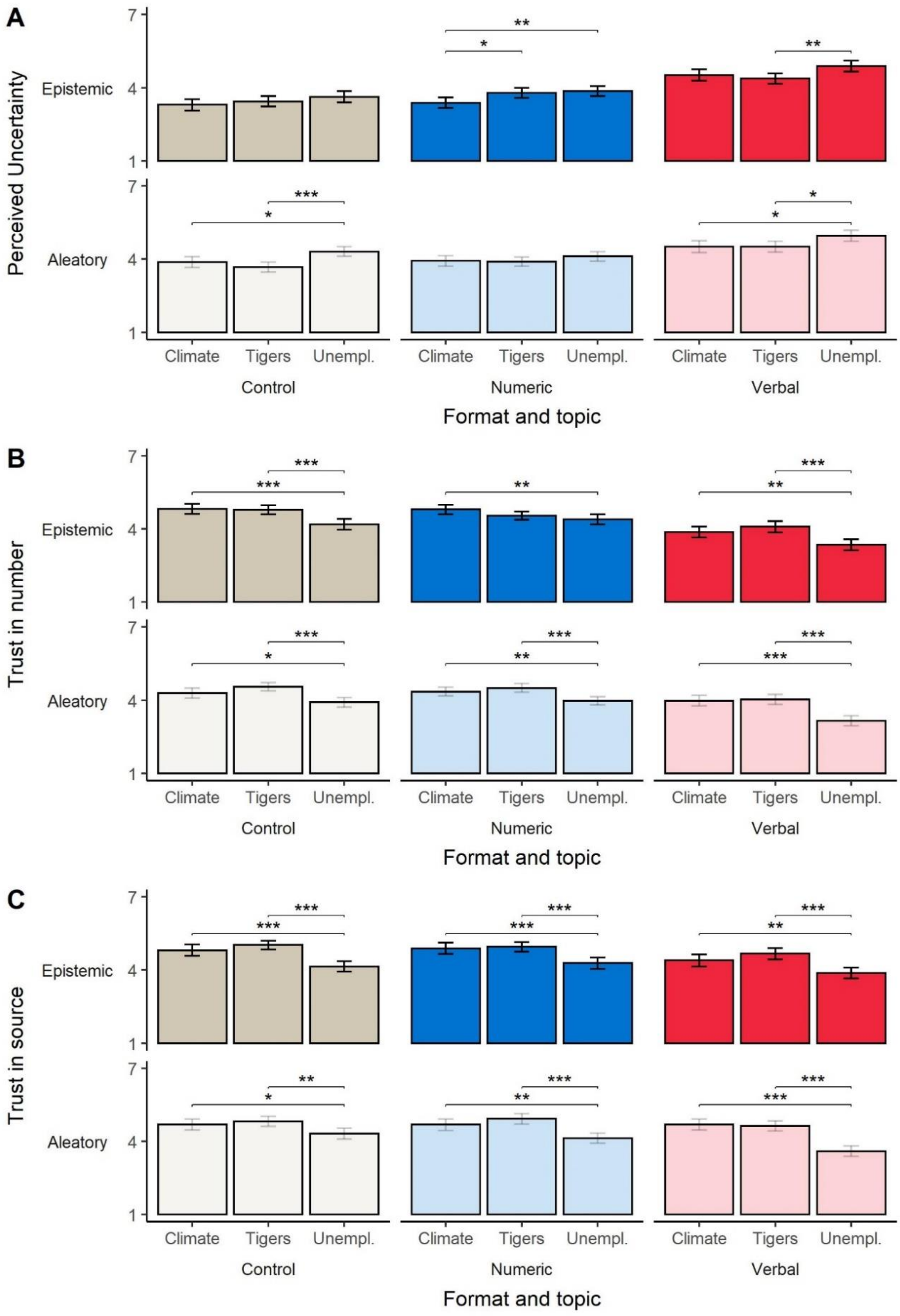

Figure S5: The effect of topic on: (A) perceived uncertainty, (B) trust in numbers, and (C) trust in source (means and 95% CI). Horizontal bars indicate a significant pairwise difference between topic conditions, based on one-way ANOVA with Tukey's post-hoc tests. *$p < .01$, **$p < .01$, ***$p < .001$.

**Appendix 7: Study 3 secondary outcomes**

In Study 3 participants also completed two additional secondary outcome items. Emotional response to the information was measured with the item: *How does the information you just read make you feel?* (responses: *negative/unhappy* (0) to *positive/happy* (10)). Reported difficulty was measured with the item: *How easy or difficult did you find to understand this number?* (*very easy* (1) to *very difficult* (7)).

Average responses across experimental conditions are reported in Supplementary Table 4. As with the analyses in the main text, we analysed responses for each topic separately with 2(type)x3(format) two-way ANOVAs.

Considering reported difficultly, we report a significant main effect of format in the climate topic condition, $F(2, 747) = 5.28$, $p = .005$, $\eta_p^2 = .014$, with numeric formats rated as more difficult to understand than the control (separate post hoc tests comparing formats within each uncertainty type indicated this pairwise difference was only significant in the epistemic condition, see Figure S6A).

We also report a significant main effect of format in the tigers topic condition, $F(2, 751) = 12.23$, $p < .001$, $\eta_p^2 = .032$, with numeric formats rated as more difficult to understand than the control and verbal uncertainty formats (see Figure S6A).

For unemployment messages we report a significant main effect of uncertainty type, $F(1, 737) = 7.95$, $p = .005$, $\eta_p^2 = .011$, but not format. Aleatory uncertainty was rated as more difficult understand than epistemic uncertainty.

Considering emotional response, we found no significant main effects or interactions (all $p > .15$; see Figure S6B).

Table S6. Study 3 secondary outcomes across experimental conditions (mean (SD))

| Conditions | | | n | Positive feelings | Difficulty |
|---|---|---|---|---|---|
| Topic | Type | Format | | | |
| Climate | Epistemic | Control | 121 | 2.62 (2.16) | 2.65 (1.53) |
| | | Numeric | 125 | 2.36 (2.38) | 3.13 (1.37) |
| | | Verbal | 126 | 2.66 (1.98) | 2.97 (1.54) |
| | Aleatory | Control | 128 | 2.52 (2.20) | 2.73 (1.28) |
| | | Numeric | 128 | 2.27 (2.10) | 3.10 (1.59) |
| | | Verbal | 125 | 2.69 (2.20) | 2.77 (1.48) |
| Tigers | Epistemic | Control | 127 | 1.61 (1.79) | 2.47 (1.31) |
| | | Numeric | 125 | 1.66 (1.78) | 3.18 (1.45) |
| | | Verbal | 124 | 1.88 (1.85) | 2.61 (1.40) |
| | Aleatory | Control | 127 | 1.60 (1.95) | 2.71 (1.46) |
| | | Numeric | 126 | 1.62 (1.94) | 3.15 (1.39) |
| | | Verbal | 128 | 2.07 (1.94) | 2.71 (1.48) |
| Unemployment | Epistemic | Control | 118 | 3.31 (2.08) | 2.59 (1.25) |
| | | Numeric | 125 | 3.26 (2.12) | 2.88 (1.34) |
| | | Verbal | 124 | 3.32 (2.00) | 2.52 (1.30) |
| | Aleatory | Control | 122 | 2.90 (2.16) | 2.95 (1.53) |
| | | Numeric | 128 | 2.98 (2.28) | 3.00 (1.43) |
| | | Verbal | 127 | 3.33 (2.12) | 2.90 (1.36) |

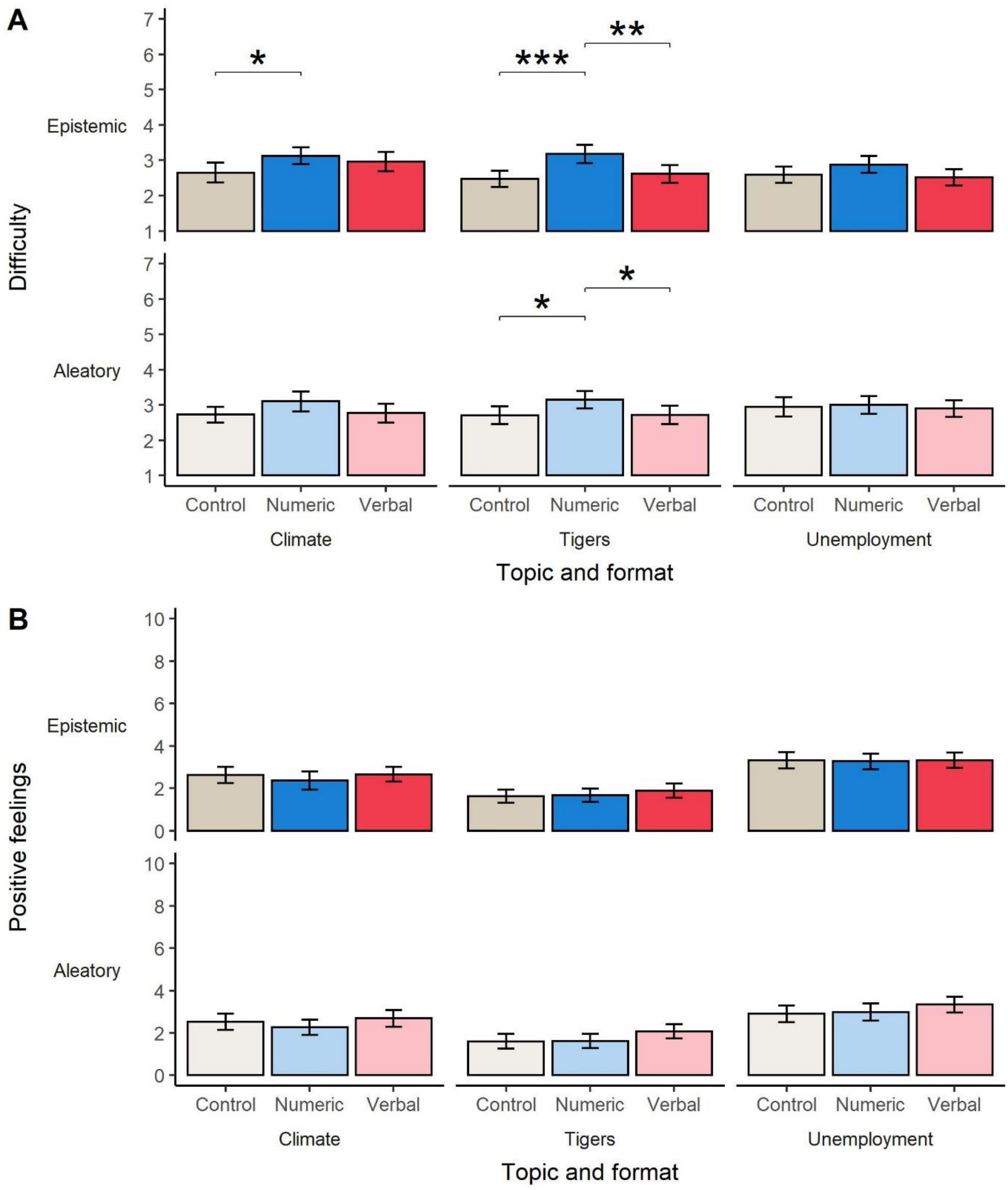

Figure S6: The effect of format on secondary outcomes: (A) reported difficulty in understanding numbers and (B) emotional response to information (means and 95% CI). Horizontal bars indicate a significant pairwise difference between format conditions, based on one-way ANOVA with Tukey's post-hoc tests. *$p < .05$, **$p < .01$, ***$p < .001$.

Use the "Insert Citation" button to add citations to this document.
